# Feasibility of Inferring Spatial Transcriptomics from Single-Cell Histological Patterns for Studying Colon Cancer Tumor Heterogeneity

**DOI:** 10.1101/2023.10.09.23296701

**Authors:** Michael Y. Fatemi, Yunrui Lu, Cyril Sharma, Eric Feng, Zarif L. Azher, Alos B. Diallo, Gokul Srinivasan, Grace M. Rosner, Kelli B. Pointer, Brock C. Christensen, Lucas A. Salas, Gregory J. Tsongalis, Scott M. Palisoul, Laurent Perreard, Fred W. Kolling, Louis J. Vaickus, Joshua J. Levy

## Abstract

**Background:** Spatial transcriptomics involves studying the spatial organization of gene expression within tissues, offering insights into the molecular diversity of tumors. While spatial gene expression is commonly amalgamated from 1-10 cells across 50-micron spots, recent methods have demonstrated the capability to disaggregate this information at subspot resolution by leveraging both expression and histological patterns. However, elucidating such information from histology alone presents a significant challenge but if solved can better permit spatial molecular analysis at cellular resolution for instances where Visium data is not available, reducing study costs. This study explores integrating single-cell histological and transcriptomic data to infer spatial mRNA expression patterns in whole slide images collected from a cohort of stage pT3 colorectal cancer patients. A cell graph neural network algorithm was developed to align histological information extracted from detected cells with single cell RNA patterns through optimal transport methods, facilitating the analysis of cellular groupings and gene relationships. This approach leveraged spot-level expression as an intermediary to co-map histological and transcriptomic information at the single-cell level.

**Results:** Our study demonstrated that single-cell transcriptional heterogeneity within a spot could be predicted from histological markers extracted from cells detected within a spot. Furthermore, our model exhibited proficiency in delineating overarching gene expression patterns across whole-slide images. This approach compared favorably to traditional patch-based computer vision methods as well as other methods which did not incorporate single cell expression during the model fitting procedures. Topological nuances of single-cell expression within a Visium spot were preserved using the developed methodology.

**Conclusion:** This innovative approach augments the resolution of spatial molecular assays utilizing histology as a sole input through synergistic co-mapping of histological and transcriptomic datasets at the single-cell level, anchored by spatial transcriptomics. While initial results are promising, they warrant rigorous validation. This includes collaborating with pathologists for precise spatial identification of distinct cell types and utilizing sophisticated assays, such as Xenium, to attain deeper subcellular insights.

## Background

Cancer warrants considerable attention given its profound impact on individuals and their families, the absence of a clear cure in many cases, and the necessity to advance technologies for prevention, early detection, and treatment selection. By the end of 2023, nearly 2 million new cancer cases and more than 600,000 cancer deaths will occur in the United States [1,2]. Colorectal cancer (CRC) persists as one of the most formidable solid tumors, with an annual incidence of approximately 150,000 new cases in the United States and a 63% 5-year survival rate [1,2]. With the shift in CRC to younger demographics and tumor metastasis being responsible for most cancer deaths, there is a pressing need for a high-fidelity screening and prognostication program [3]. The treasure trove of imaging and genomics information provided by nascent molecular assays and informatics techniques has the potential to inform more effective, targeted treatment options by revealing novel prognostic biomarkers.

The pathological TNM-staging system (pTNM) is the most predictive factor for the risk of recurrence and prognosis [4–7]. Cancer staging is broadly characterized by assessing local invasiveness (T-stage), followed by the extent of lymph node involvement (N-stage), then the presence of metastasis to distant sites. Successful extraction of lymph nodes at the time of resection is crucial for establishing the N-stage, which primarily informs the risk of recurrence [6]. A higher yield of lymph nodes during surgery is often associated with favorable outcomes at the expense of potential morbidity; as such, the recommended lymph node yield is set at 12 [6,8]. Yet, one recent population study found that only 37% of such assessments extract this number, underscoring significant implications for accurate risk assessment and selection of optimal therapies [4]. For example, the usage of neoadjuvant chemotherapy is typically considered for stage III patients with proficient mismatch repair (MMR) status. As there are instances where the presence of metastasis is uncertain, thereby impacting the fidelity of prognostic assessments, informatics tools that can study the spatial biology of the tumor at the primary site can infer missing staging information (e.g., lymph node stage) or identify risk factors independent of pTNM staging relevant which provide additional predictive value for the recurrence risk assessment.

Tumor Infiltrating Lymphocytes (TIL) play a crucial role in understanding and modulating the Tumor Microenvironment (TME) and Tumor Immune Microenvironment (TIME) [9]. The TME consists of malignant and benign cells, blood vessels, and extracellular matrix, interconnected through complex communication via cytokine recruitment factors [9]. Recent studies highlight the importance of immune infiltrates, such as T cells, B cells, NK cells, and monocyte/lymphocyte cells, and their distribution, density, and relationships in mounting an effective anti-tumor response. Microsatellite Instability (MSI) status also influences this response [10]. For example, high levels of cytotoxic T cells within the tumor may indicate immune exhaustion, while increased immune cell density can suggest a favorable prognosis [11]. Understanding the molecular changes and spatial arrangements associated with colon cancer metastasis is still incomplete. Nonetheless, several digital pathology assays have incorporated existing findings to complement pTNM staging and serve as independent risk factors for recurrence. These assays include: 1) Immunoscore, which measures the density of cytotoxic T-cells at the tumor’s invasive margin and inside the tumor [12], 2) CDX2, an epithelial marker of pluripotency indicating the tumor’s ability to bypass immune response and growth inhibition checkpoints [13–15], and 3) circulating tumor DNA, such as mutations in the Vascular Endothelial Growth Factor (VEGF) pathway [16–18]. While these assays are predictive of recurrence risk, they provide only a limited perspective on tumor metastasis phenomenology.

Spatial omics technologies, like 10x Genomics Spatial Transcriptomics (ST) or GeoMX Digital Spatial Profiling (DSP), have facilitated the simultaneous analysis of multiple biomarkers, including the whole transcriptome, with remarkable spatial resolution [19–22]. These technologies have been applied to further characterize TIL subpopulations in TME. However, their clinical utility is limited due to high costs, low throughput, and limited reproducibility. In previous work, we demonstrated the feasibility of utilizing machine learning algorithms to extract TIL and spatial biology information from Hematoxylin and Eosin (H&E) stains. This can be a cost-effective and high-throughput digital biomarker that could be employed prospectively as an adjunct test similar to Immunoscore for recurrence risk assessment [23–25]. We found that careful selection of algorithms is crucial to capture molecular alterations and pathways reflective of histomorphological changes or large-scale tissue architecture changes [26,27].

Nevertheless, the resolution of these findings is currently restricted to the available resolution of Visium spots, typically around 50 microns, which aggregates expression data across a small number of cells (1-10 cells). Incorporating single-cell information, captured through the new Chromium Flex technology, can enable further characterization of spatial cellular heterogeneity to enhance the resolution of the Visium data. Recent advancements in 10x profiling technologies, including Chromium Flex and CytAssist assays, enable the profiling of single-cell transcriptomics (scRNASeq) on serial sections of formalin-fixed paraffin-embedded (FFPE) tissue. By utilizing automated staining devices and advanced imaging techniques prior to conducting Visium spatial transcriptomics (ST) assays, the CytAssist technology significantly improves workflow that allow for higher resolution whole slide images (WSI) to be collected. Pairing this high-resolution slide imaging information with ST and serial section scRNASeq has the potential to enhance the capacity to perform spatial assessments at single-cell resolution on external study cohorts.

While several technologies have been developed to increase the resolution of Visium data, such algorithms require both ST and histological information and do not operate on tissue images alone. Previous studies have made attempts to infer single-cell RNA sequencing (scRNA-seq) data from breast cancer tissue slide sections, improving the resolution of the data and enabling the identification of different cell types within the tissue [28]. Other studies have made attempts to infer Visium ST expression patterns aggregated across several cells per spot using image classification techniques with some domain-specific adaptations. For example, recent studies have trained DenseNet-121 and InceptionV3 models to predict gene expression [29,30], and another work used a custom convolutional layer along with a graph attention network and transformer model to share information between Visium spots [25]. While the Visium platform primarily provides low-resolution, aggregated expression measurements across cells contained within a 50-micron spot [31,32], single-cell analyses offer a more comprehensive view of cellular heterogeneity. One study attempts to reconstruct false zero-counts in cell-level omics data by training a masked autoencoder over known cell data [33], and other recent efforts have been made to directly infer single-cell transcriptomes from whole slide images by collecting information from single cells in serial sections following H&E staining, yet lacked ST data to guide the co-registration of the single cell data to the slide [34].

The primary goal of this study is to enhance the predictive capability of algorithms that infer spatial transcriptomics (ST) data solely from histology images, capturing single-cell heterogeneity within a spot and their aggregate spot-level expression. To achieve this, we combine the precise locations of individual cells, as identified in whole cell images, with the granular data from single-cell RNA sequencing (scRNA-seq). This approach integrates histological details from localized nuclei within and around Visium spots with corresponding scRNA-seq profiles mapped to the same spots. By seamlessly merging these datasets, our framework stands poised to extract richer molecular insights from cells, facilitating a more accurate prediction of both Visium ST and individual cell information.

Through feature engineering methods, we develop attribution methods to examine the structural organizations of cells that are most correlated with the expression of specific genes. This can contribute to a better understanding of the dynamics of the tumor-immune microenvironment and potentially aid in developing prognostic tools for the stage and aggressiveness of colorectal tumors. In this paper, we compare the accuracy of methods that use cells as features with conventional computer vision methods featured in our previous work. It is important to emphasize that this study does not claim to infer scRNASeq data at specific locations of individual cells. Rather, we demonstrate the ability to leverage single-cell information to enhance the expression prediction at Visium spots on held-out tissue slides. This research establishes a foundational workflow and conceptual framework for the future inference of such information.

## Results

### Overview of Cells2RNA Framework: Bridging Histological Patterns with Single-Cell Expression

Cells2RNA was crafted to infer single-cell expression insights from discernible histological patterns in instances where spatial transcriptomics and single-cell data might be lacking (**Figure 1**). The crux of the challenge lies in deducing single-cell nuances solely from histological patterns surrounding pinpointed cells (**Figure 2A**). Historically, prior research has been limited to interpreting aggregated spot-level data. Yet, when disaggregated to the individual cell level, a richer tapestry of heterogeneity emerges, which becomes our focal point for inference. This innovation holds the potential to augment findings across expansive cohorts without the constraints of traditional spatial analyses that heavily rely on Visium data. The goal of this study is to derive molecular insights paralleling the depth of Visium-based investigations, but strictly from histological imaging.

**Figure 1:**
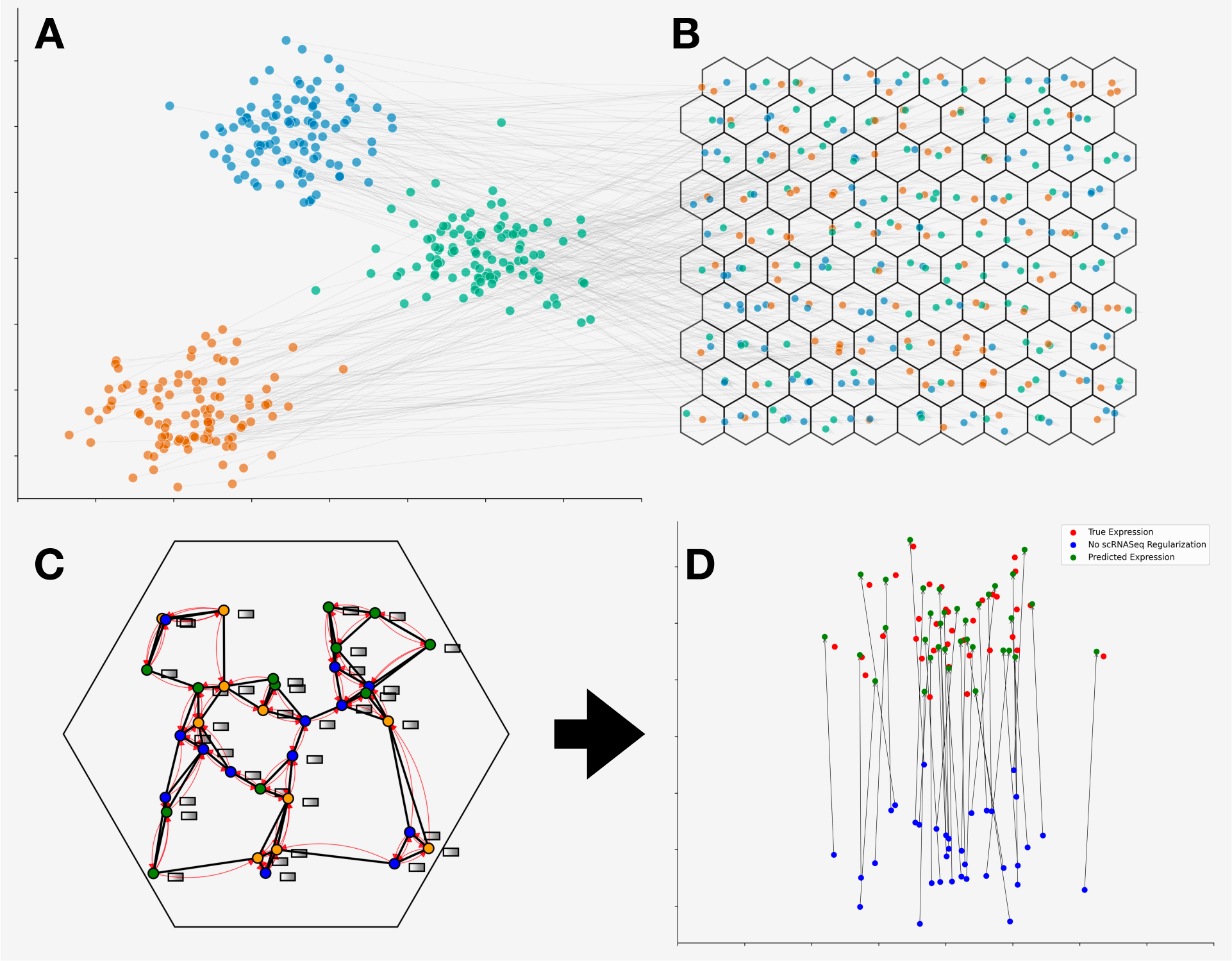
Overview of Cell2RNA’s Co-Mapping Approach. (A) Low-dimensional visualization of single-cell RNA profiles, clusters indicating cell-type. (B) Spatial layout of identified cells across the tissue slide (assignment to spots represented by hexagons), color-coded by distinct gene expression patterns mapped from single cell profiles featured in (A). (C) In-depth view of cells located within a specific Visium spot, illustrating connectivity and cell relationships. Expression-related histological features, represented by grey rectangles, are shared among neighboring cells through red curves via a graph neural network. (D) A side-by-side low-dimensional comparison of scRNASeq profiles for a representative Visium spot: actual expression (red), model-predicted expression using the co-mapping training approach (green), and expression prediction without co-mapping training (blue).

**Figure 2:**
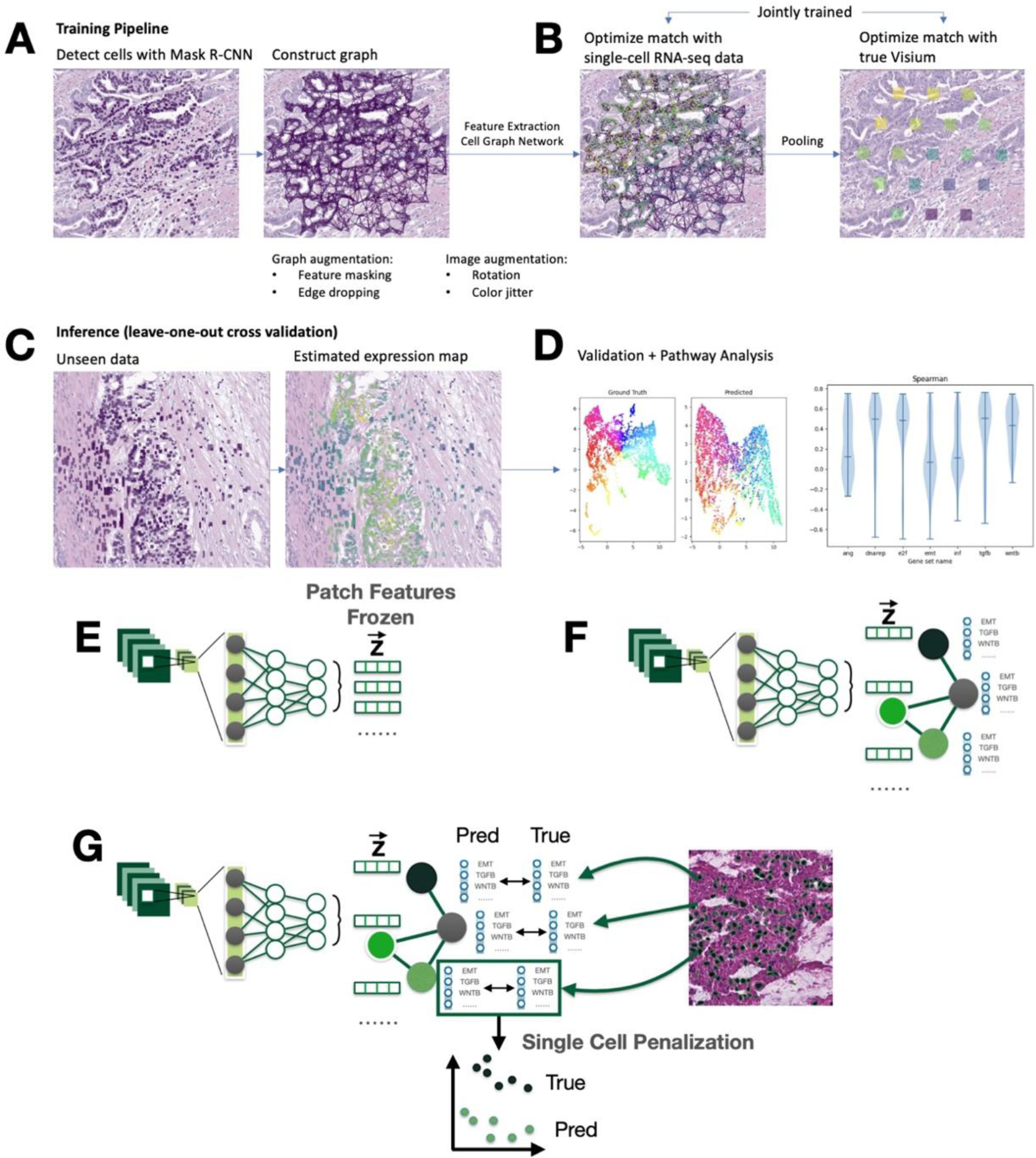
Schematic Representation of the Neural Network Workflow for Single-Cell Analysis. During the training phase, (A) a pre-trained Mask R-CNN model is applied to raw histology images to detect individual cells, after which a 6-nearest neighbors graph is constructed for the detected cells. (B) Features for each cell are extracted using a ResNet-50 neural network, and the aggregation of neighboring cell information is modeled using a Graph Attention Network (GAT). For each Visium spot, the node features are aggregated using sum pooling. (C) Pre-pooled node values are jointly optimized against single-cell RNA-sequencing (scRNA-seq) data, and (D) pooled Visium spot predictions are optimized against the corresponding ground truth data, using a mean-squared-error loss computed across log-transformed counts. (E)-(G) Visual description of neural network architectures and penalizations employed: (E) a two-stage neural network comprising a feature extraction stage and a prediction stage, this was not used in this work, (F) an end-to-end neural network encompassing the entire process from cell detection to feature extraction, graph convolutions and prediction, utilized in this study, and (G) the incorporation of single-cell-level penalties into the loss function to enforce consistent predictions with scRNA-seq data.

Central to our approach is a co-mapping methodology. Here, histological patterns detected at the cellular level are intricately aligned with single-cell expression data (**Figure 1A**). Spatial transcriptomics serves as a vital intermediary in this process: during training, it not only maps single-cell RNASeq data to corresponding Visium spots (**Figure 1B**) where cells are located but also acts as a crucial inference target for the expression-centric histological attributes derived from these located cells.

To accomplish this, Cell Graph Neural Networks (CGNNs) are employed (**Figure 2B,E,F**). They efficiently extract the essence of histological features from located cells within Visium spots (**Figure 1C**). These features are updated during model training through simultaneous harmonization with scRNASeq data via optimal transport methodologies (**Figures 1D,2G**), forging a cohesive link between cellular histological characteristics and their intricate gene expressions.

The culmination of this methodological synergy is a framework where the expression profiles predicted from detected cells align with the broader single-cell expression landscape. Although this alignment might not be perfect, it closely mirrors genuine single-cell expression dynamics within each Visium spot. Thus, our anticipation is twofold: the accurate convergence of aggregated inferred single-cell expression to spot-level benchmarks and a holistic reflection of single-cell expression diversity within the spot when applied to external, held-out slides (**Figure 2C,D**).

Using Visium and paired 40X resolution whole slide imaging from a cohort of nine stage pT3 colorectal patients (see section “Data Collection and Preprocessing”), the co-mapping technique was benchmarked against patch-level models (Inceptionv3) and other CGNNs that utilize alternative information extraction methods. We assessed their performance based on predicting spot-level expression, capturing cellular heterogeneity within spots (using Wasserstein distance), maintaining tissue architectural relationships, and conducting a pathway analysis. A detailed examination of these methodologies and comparisons is provided in the Methods section.

### Model Comparison

Overall, models have strong performance– selecting the top CGNN model per gene resulted in an AUROC of 0.8138 ± 0.0069 and Spearman’s statistic of 0.5724 ± 0.0133 (**Table 1**). However, across all experiments, model performances did not appear significantly different from each other, though we noticed several important trends (**Figure 3,4**). CGNN models performed on par with the Inception model (AUC=0.8204 ± 0.0073). The most predictive cell-based model had an AUROC of 0.8093 ± 0.0083, similar to the InceptionV3 model’s AUROC interval of 0.8204 ± 0.0073, which leveraged additional information beyond the cell’s immediate neighborhood and may have also benefited from the built-in structural feature extraction of CNNs. There was high agreement in top-performing genes between CGNN methods using graph contrastive learning or single-cell penalization as compared to a CGNN with no penalization/pretraining (**Supplementary Figure 1, Supplementary Table 1**). Notable genes with high predictive performance include TMSB4X, which encodes an actin-sequestering protein vital for actin polymerization, cell proliferation, migration, differentiation, and bypasses X inactivation with a homolog on chromosome Y; and ELF3, which regulates the inflammatory response (**Supplementary Table 1**).

**Figure 3:**
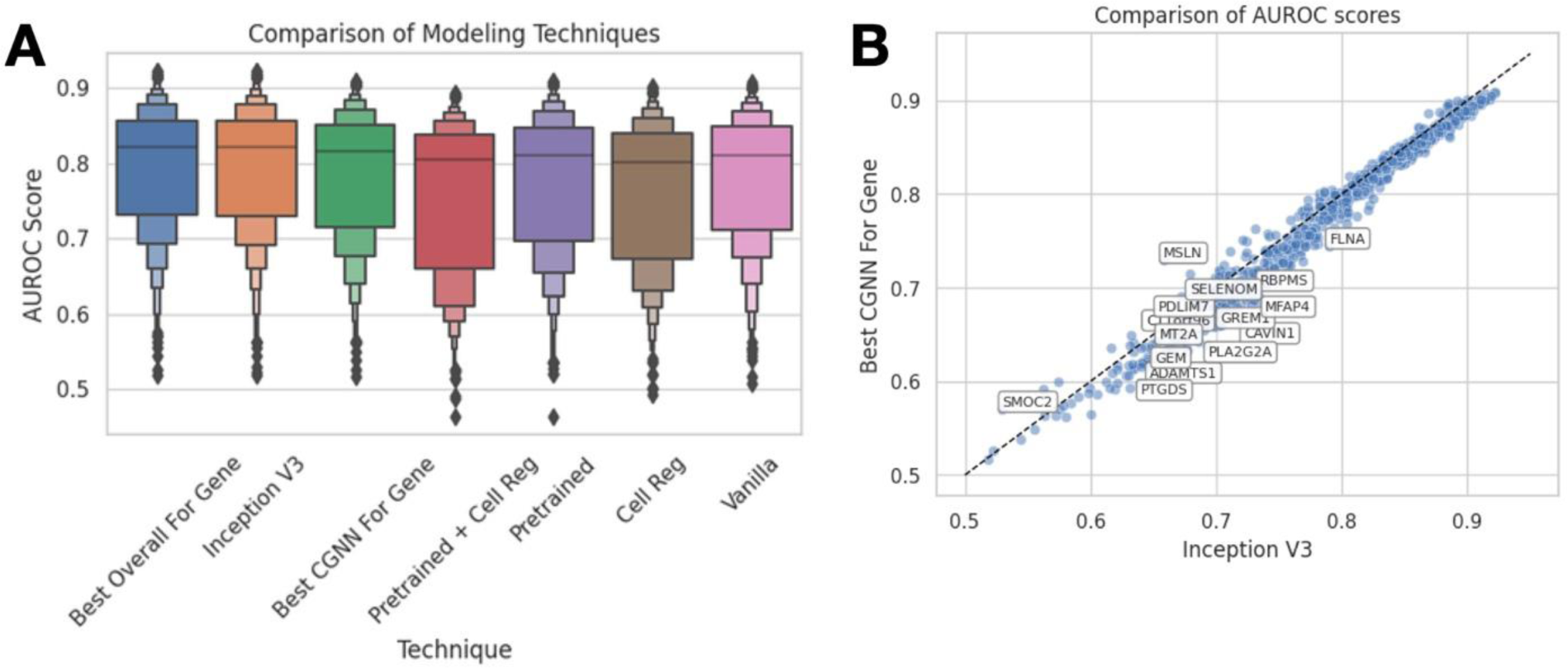
Performance comparison between methods. **A)** Boxplot of AUROC scores from each method; **B)** comparison of AUROC for best CGNN and CNN for each gene

**Figure 4:**
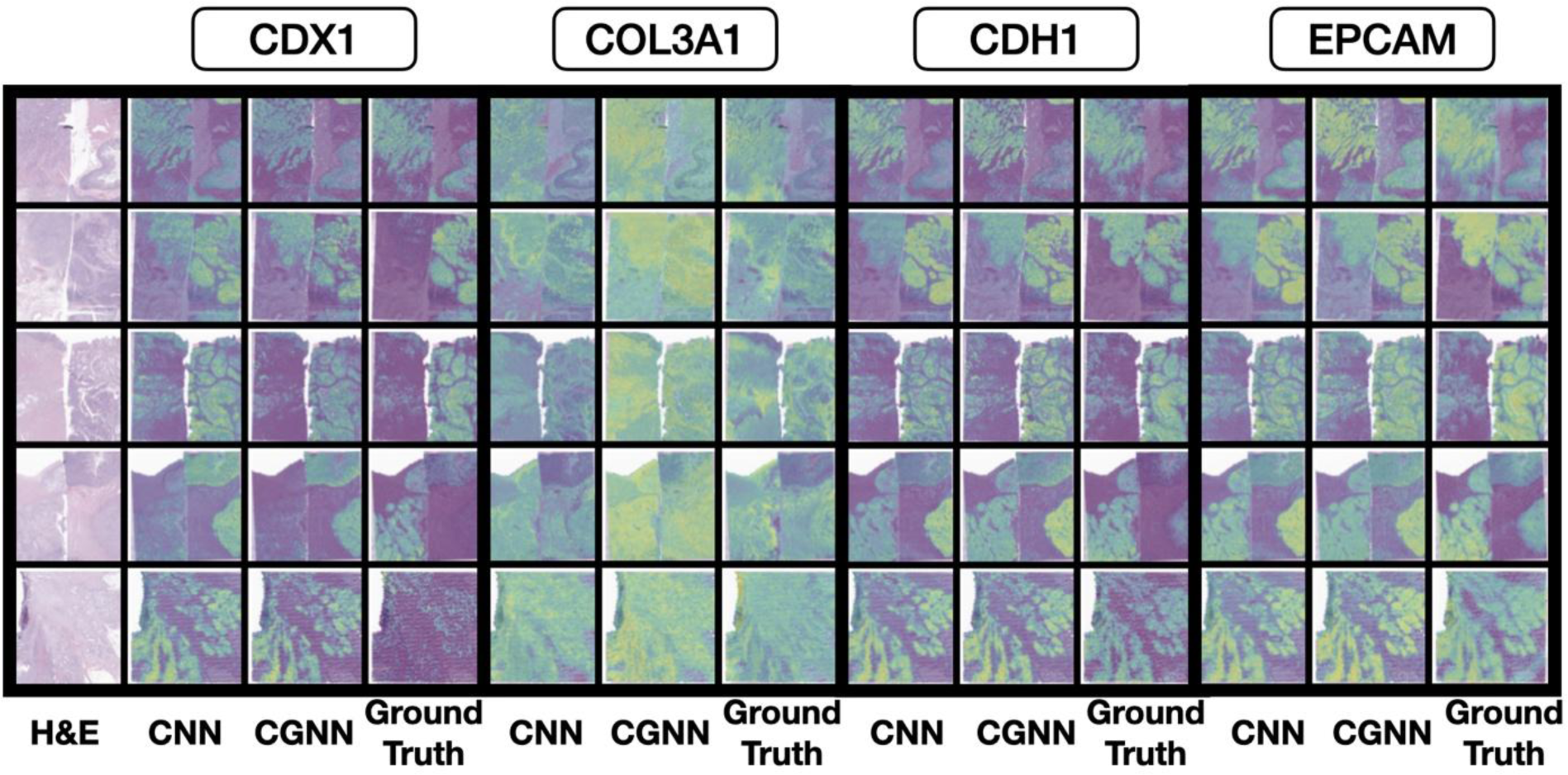
Predicted expression for various genes. CNN, CGNN, compared to ground truth for genes *CDX1, COL3A1, CDH1* and *EPCAM* across sections from all nine patients

**Table 1:**
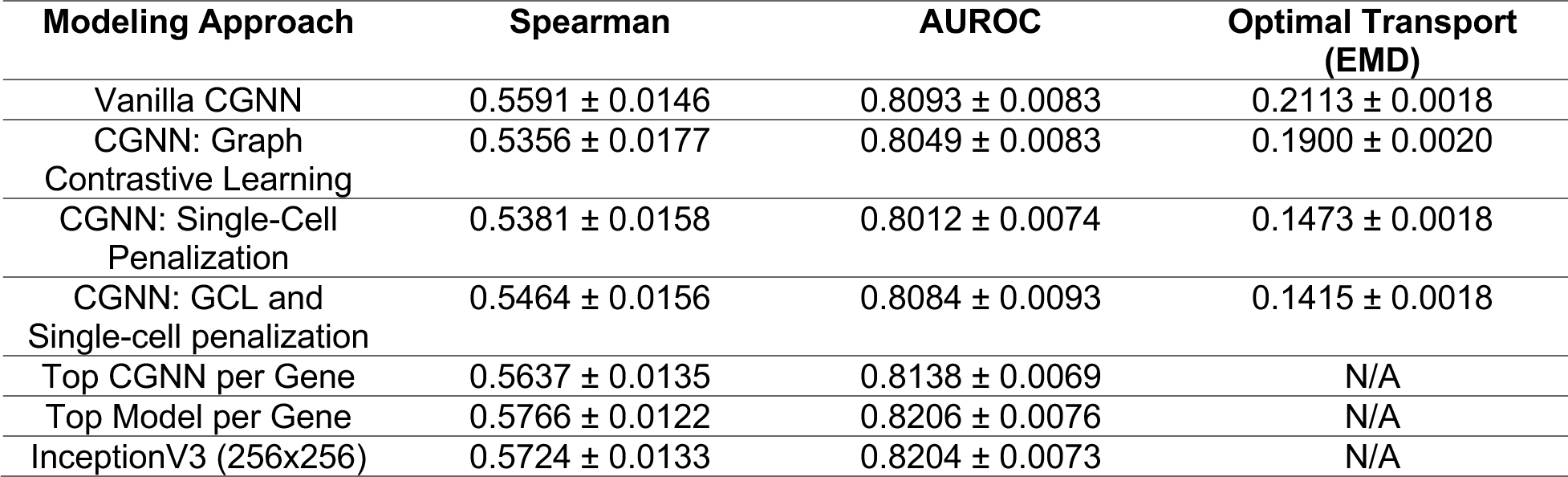
Comparison of model performance. Aggregate AUROC is calculated as the median AUROC across genes. Gene-level AUROC is calculated as the mean across cross-validation folds.

| Modeling Approach | Spearman | AUROC | Optimal Transport (EMD) |
| --- | --- | --- | --- |
| Vanilla CGNN | $0.5591 \pm 0.0146$ | $0.8093 \pm 0.0083$ | $0.2113 \pm 0.0018$ |
| CGNN: Graph Contrastive Learning | $0.5356 \pm 0.0177$ | $0.8049 \pm 0.0083$ | $0.1900 \pm 0.0020$ |
| CGNN: Single-Cell Penalization | $0.5381 \pm 0.0158$ | $0.8012 \pm 0.0074$ | $0.1473 \pm 0.0018$ |
| CGNN: GCL and Single-cell penalization | $0.5464 \pm 0.0156$ | $0.8084 \pm 0.0093$ | $0.1415 \pm 0.0018$ |
| Top CGNN per Gene | $0.5637 \pm 0.0135$ | $0.8138 \pm 0.0069$ | N/A |
| Top Model per Gene | $0.5766 \pm 0.0122$ | $0.8206 \pm 0.0076$ | N/A |
| InceptionV3 (256x256) | $0.5724 \pm 0.0133$ | $0.8204 \pm 0.0073$ | N/A |

### Single-Cell Attribution Maps Point to Spatial Cellular Heterogeneity

Single-cell regularization was able to significantly improve the alignment of cellular information extracted from located cells within held-out slides with their corresponding single-cell profiles, as measured by the Earth Mover’s (Wasserstein) distance between cells assigned to spots using Tangram and their closest detected matches (EMD=0.1415 ± 0.0018 with penalization, 0.2113 ± 0.0018 without penalization). This improvement does not negatively impact AUROC. Cells were embedded using UMAP based on the ground truth and predicted expression, with and without penalization with scRNASeq. Visual inspection of these UMAP embeddings confirmed the quantitative results of differences in EMD (**Supplementary Figure 2**), that single-cell penalization causes node-level predicted expression from cellular histomorphology for genes to more closely resemble the distribution of single-cell data assigned to the Visium spot with Tangram.

An optimal transport approach matched single-cell profiles between the predicted and ground truth. We then assessed the correlation between individual cells for specific genes based on their performance on the Visium spot level. Overall, more than 80% of the genes exhibited a positive correlation between ground truth and predicted single-cell expression when single-cell regularization was employed, compared to around 20-30% of the genes without such regularization was not used (**Supplementary Figure 3**). Examining inferred cell-level expression before the final pooling layer makes the source of our network’s predicted gene expression evident. As illustrated in **Figure 5E-G**, we juxtapose the predicted level of EPCAM expression for each cell against ground truth data from a Visium assay. Notably, the Visium assay offers aggregate expression measurements spanning a broader region. Our model’s predictions and the ground truth at cellular resolution are visually consistent (**Figure 5A-D**), further corroborating with both the high accuracy results reported from the previous section as well as the lower EMD reported through single-cell penalization.

**Figure 5:**
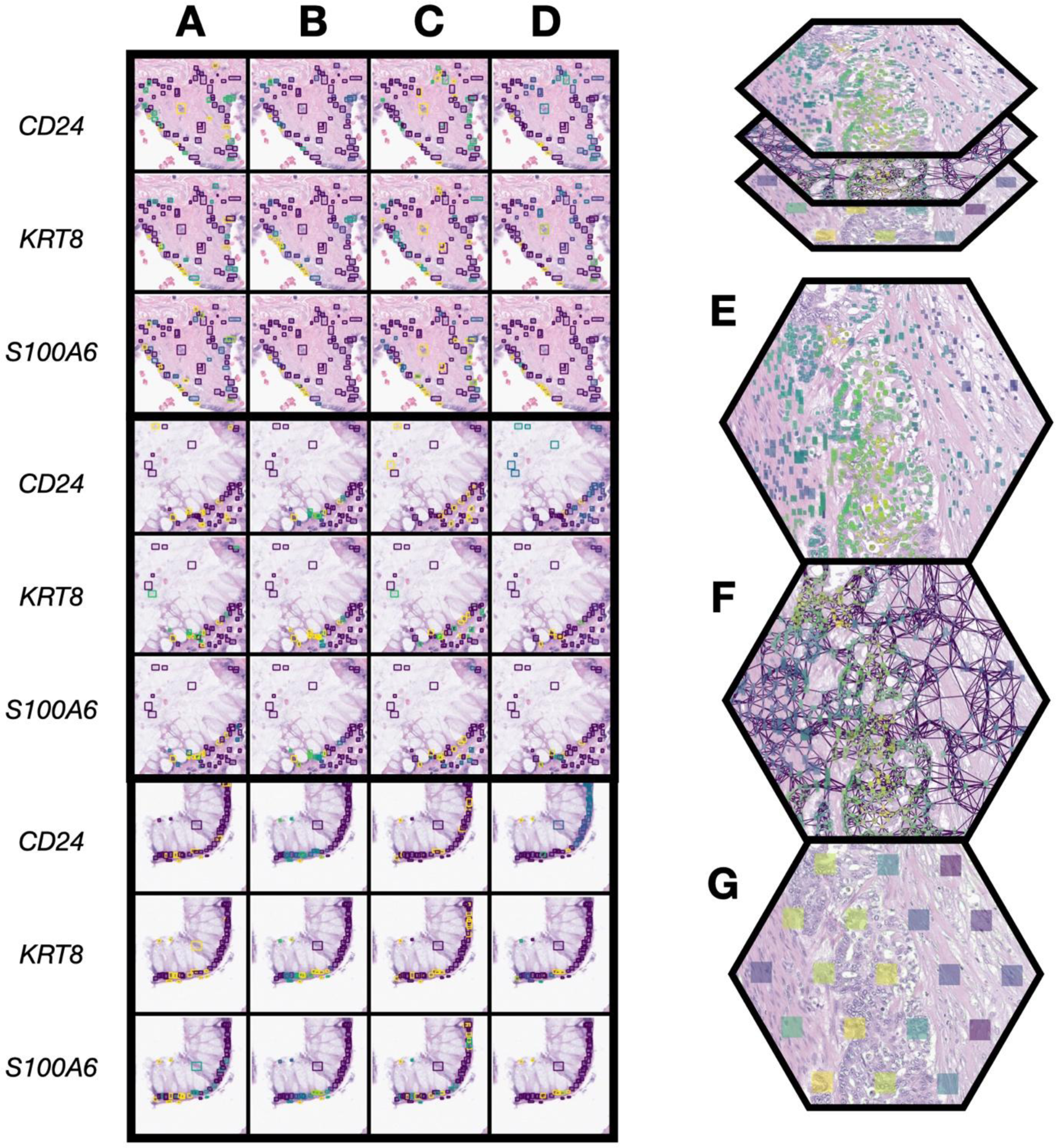
Alignment of True and Predicted Single-Cell and Visium-Spot Level Expression on a Histological Section. The figure illustrates the relationship between true and predicted single-cell expression on a histological section for genes CD24, KRT8, and S100A6. Ground truth expression was assigned to each cell using optimal transport based on comparing predicted and actual expressions rather than their spatial arrangement. A) and C) display the ground truth of single-cell expression with and without single-cell regularization, respectively. B) and D) visualize the respective predicted single-cell expressions. Progressing from individual cellular predictions to a broader view, D)-G) detail the transition through EPCAM expression: from predicted cell-level expression in D) to an overarching cell graph across multiple Visium spots in E) and concluding with spot-level Visium expression in G).

Cell-level attribution (i.e., inferred expression) maps mirror tissue architecture. A closer inspection of a tissue section reveals patterns, such as cells exhibiting high expression juxtaposed with a cluster of low-expression cells. Such patterns potentially highlight activities conforming to the underlying tissue structure. The structural coherence of granular cell-level information likely stems from our regularization strategies, compelling the neural networks to internalize a general representation of cell graphs and the sheer volume of available data. In instances where Visium spots exhibit diverse expression levels yet have similar cell graphs, the graph neural network may pinpoint certain cells that act as primary influencers or representatives of the expression within that spot (i.e., have markedly similar/different expression for specific genes). These specific cells could either epitomize the overall expression profile or significantly sway the expression heterogeneity at that location. Further validation is necessary to corroborate these findings, and this will be a central focus of our future research.

### Topological Consistency of Inferred Expression Patterns

Across all capture areas, predicted spot level expression clustered similarly to the true expression as the relative positioning between the true and predicted clusters in the UMAP plots was preserved (**Figure 6**). However, there were notable differences. Overlaying the clusters assigned to ground truth embeddings over the predicted expression embeddings, we found that clusters were less separated, more connected, clumped, and fuzzier than the ground truth. Nonetheless, overlaying cluster assignments across the whole slide image demonstrates the ability of these models to derive expression signatures that can delineate key histological architectures that will be the subject of inquiry in future work.

**Figure 6:**
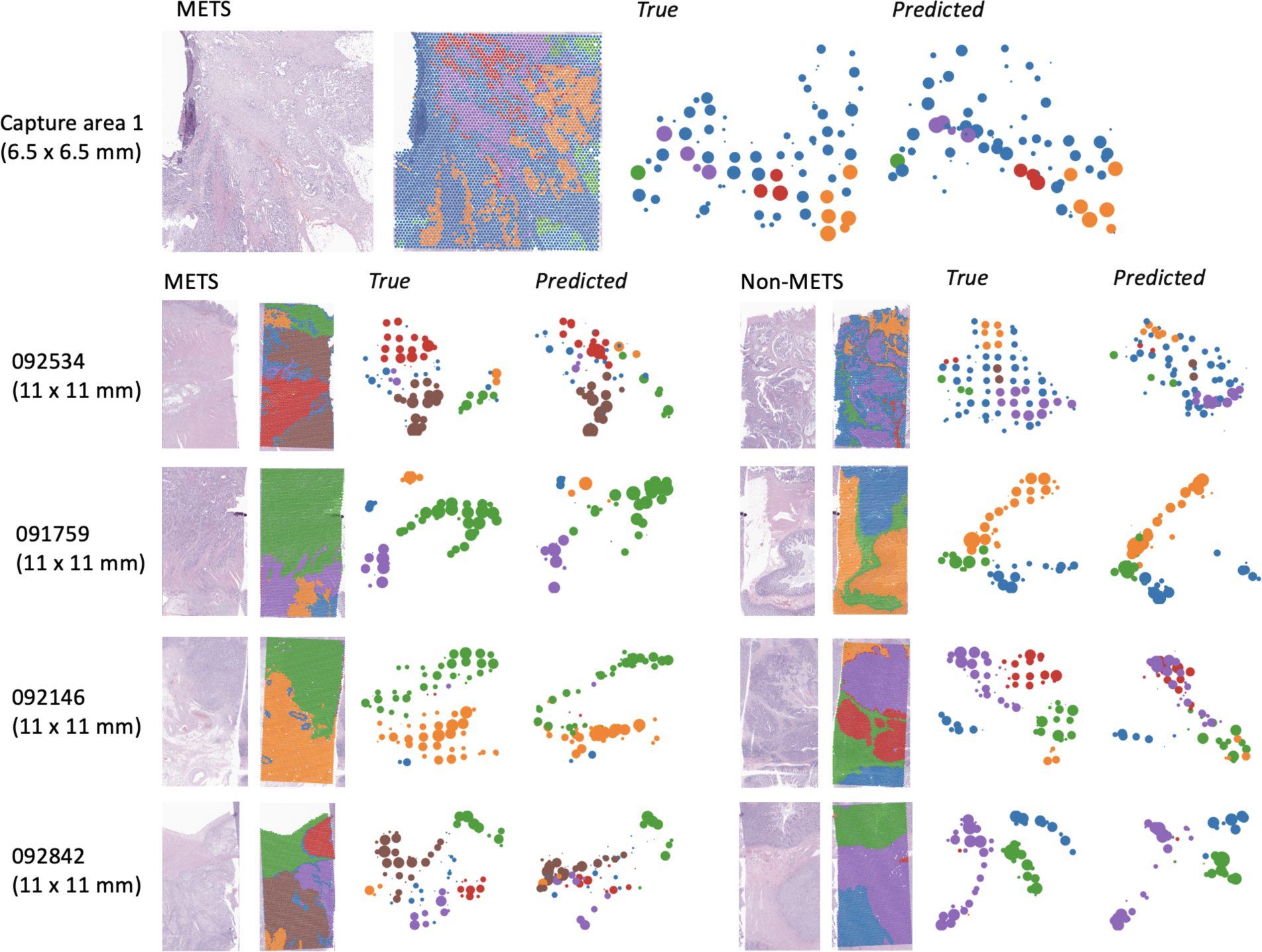
UMAP embeddings of tissue slides from selected capture areas, color-coded by HDBSCAN clusters. Comparisons include CGNN, CGNN with single-cell penalization, and patch-based methods against the ground truth. Clusters derived from the ground truth are overlaid on the slides for context.

### Pathway Analysis

To compare performance across several potential prediction targets, we selected pathways from MSigDB’s Hallmark Gene Sets [35,36] and reported the average AUC for genes from these sets. Across all modeling approaches, genes involved in DNA repair and E2F targets were predicted with higher performance as compared to other molecular pathways (**Supplementary Figure 4**). Dysregulation of DNA repair can impact the acceleration of tumor progression [37], and therefore accurately detecting the presence of relevant genes may be useful in the prognostication of the speed of progression. We did notice that for some pathways, e.g., *Epithelial to Mesenchymal Transition*, penalizing by single-cell expression did lead to some loss of performance in distinguishing these molecular signatures (**Supplementary Figure 4**).

We performed a pathway analysis by subsetting the top 10% of genes (100 out of 1000) per modeling approach for further analysis using the Enrichr software/database [38–41]. The top ten statistically-significant pathways (from the Elsevier pathway collection), divided by each modeling approach, are shown in **Supplementary Table 2**. We also found that the *WNT in Epithelial to Mesenchymal Transition in Cancer* pathway, a chief contributor to the migration and metastasis of cancer cells, and several pathways associated with *desmosome assembly* (which regulate intercellular adhesion between metastasizing cells) were among the top ten most statistically significant gene sets detected in all four techniques, and *EPCAM in Cancer Cell Motility and Proliferation* is a statistically significant gene set in all four techniques. The *WNT in Epithelial to Mesenchymal Transition in Cancer* pathway has an AUROC of 0.8686 ± 0.0273 for the Inception model and 0.8638 ± 0.0238 for the “vanilla” cell graph model.

## Discussion

In this research, our primary objective was to draw inferences about spatial mRNA expression patterns from whole slide images (WSI), specifically by fusing single-cell histological and transcriptomic data. One of the key advantages of deriving such spatial information from routine stains is the potential to substantially reduce the costs associated with understanding the tumor immune microenvironment (TIME). Instead of relying on expensive spatial molecular staining for multiple targets, this technique offers an economical avenue for spatial molecular assessment, which can subsequently aid in the risk evaluation of recurrence. It is becoming clear that the spatial positioning, functional status, and mere presence of immune cells within TIME are crucial determinants of the tumor’s immune response. We pioneered a cell graph neural network algorithm in response to this understanding. This was designed to meticulously analyze cell structures within WSI and associate them with specific gene expressions. The development of this algorithm highlights the viability of utilizing spatial transcriptomics as a rich pretraining source, using scRNASeq to guide single-cell level interpretations that could benefit from graph-based representations. As we merged scRNASeq data with detailed histological imaging, we also introduced attribution methods that aspire to clarify the expression patterns evident in Visium spots. Even though these methods are still evolving and need further optimization and comprehensive validation, they usher in a promising avenue to understand the nuanced relationship between spatial positioning and gene expression. Importantly, our work underscores the potential of harnessing neural networks to derive molecular insights directly from tissue histology without necessitating additional assays, even though such assays played a critical role in training our models.

Our study revealed that by considering cells’ histomorphology and spatial relationships, we could effectively predict gene expression patterns across whole slide images. In some instances, these approaches outperformed traditional patch-based computer vision methods that rely on regression models using cropped images around each Visium spot. However, the predictive capacity of these approaches was found to be similar to patch-based methods, which is not surprising considering that the cells are contained within these patches and should present some loss of information. By explicitly incorporating cells as nested observations, attribution methods enabled the identification of structural cell organizations that exhibited the strongest correlation with the expression of specific genes. This finding has the potential to enhance our understanding of the tumor-immune microenvironment dynamics in future work.

### Comparison of Cell-Level Approach to Patch-Based Methods

The performance of the CNN model does not surpass that of the cell-based approaches. Interestingly, our basic cell model, devoid of any augmentations or pretraining, demonstrates a bootstrapped AUROC confidence interval overlapping with that of the Inception model. This indicates that, even when operating with potentially less diverse information like the extracellular matrix and connective tissue, the cell-based model remains competitive against its CNN counterpart. We posit that patch-based CNNs have an inherent advantage due to their richer input dataset and innate spatial reasoning granted by 2D convolutional kernels. In contrast, graph neural networks (GNNs) rely exclusively on adjacency matrices for spatial information. The crux here is the trade-off between robustness and interpretability. Although CNN may show a slight performance advantage, its insights are limited to single-pixel attributions, neglecting the broader scope of cell-cell interactions. Conversely, the GNN model offers superior explainability, permitting direct visualization of pivotal cell-cell interactions for particular genes, with tools like Captum, GNNExplainer, and topological methods that can be used to decipher important structural motifs.

### Impact of Single-Cell Penalization

Several of our experiments, including single-cell penalization and contrastive pretraining, showed minimal influence on the final outcome. This lack of influence indicates that employing single-cell penalization can shed light on the spatial nuances of cellular disparities without compromising performance, emphasizing the significance of cellular phenomena and connectivity. We believe this is due to the large dataset size (more than 60,000 Visium spots), which may mitigate the need or potential benefit of pretraining. Additionally, although we hoped that single-cell penalization would improve the model’s robustness (by grounding predictions in real single-cell RNA quantification), the penalization provided modest performance gains over methods that did not employ this penalization. This modest gain suggests that models may produce the same optimum regardless of the intermediate feature values (i.e., cell-level predictions). However, we do note that single-cell penalization causes cell-level values to be more aligned with true single-cell data, which may indicate potential future applications in deconvolution at single-cell resolution pending further validation. Cell regularization seems to have a slight negative impact across all pathways, but this might be expected since it more directly impacts node predictions rather than graph predictions. Without this regularization, the model is more directly trained towards Visium alignment, on which it is validated. However, with the regularization, we notice that agreement between node-level predictions and single-cell profiles improves (as measured by a decrease in mean-squared error). Therefore, we propose that this method would be useful in exploring the potential for graph neural networks in cell deconvolution.

### Revisiting Topological Consistency and Intermediate Histologically-Associated Molecular States

We discovered that although the predicted expression patterns mirrored the essential topological relationships tied to specific histological structures, they were more intertwined compared to the true expression, resulting in less pronounced clustering. Such mixed clustering might suggest that these clusters signify different degrees of cellular activity for various phenomena. It seems easier for machine learning models to distinguish between low and high activity levels, but interpolating intermediate levels of activity poses a challenge from a visual standpoint. In some instances, while the ground truth UMAP expression plot showed nuanced expression, the predicted expression indicated centralized expression around specific profiles. This implies a potential loss of intricate genetic interrelations, which is expected when deducing genomic data mainly from visual information in histology. This nuanced loss in the distinctiveness of the models’ topological embeddings could indicate the limitation of extracting molecular data from histology alone, suggesting richer molecular details exist beyond just histology. A potential enhancement might involve using single-cell expression profiles to better guide the model toward a more accurate gene distribution representation. Nevertheless, overall, the model’s predictions are topologically in line with the ground truth. Areas of tissue with similar ground truth measurements also exhibit similar predicted expressions.

### Reflections on Pathway Analysis

The *WNT in Epithelial to Mesenchymal Transition in Cancer* and *EPCAM in Cancer Cell Motility and Proliferation* were notable pathways from the results section. Wnt/β-catenin signaling is implicated in cell differentiation and proliferation and has been implicated in increasing the number of “stem-like” cells in a tumor [42]. EPCAM is responsible for modulating epithelial cell adhesion, and - while having conflicting trends in recent research - can result in adhesive and migratory cell activity, potentially impacting the potential for metastasis [43].

### Immunological Considerations

Our approach to unveil single-cell heterogeneity from whole slide images through alignment with single-cell expression bears several important immunological implications. First, the spatial arrangement of immune cells not only influences processes governing the anti-tumoral response but may offer insights as to the efficacy of immunotherapies including checkpoint inhibitors which has been a timely subject of inquiry [44,45]. Deciphering the spatial make-up may also further reveal how tumors can establish immunosuppresive environments or contribute to an immune exhaustion phenotype [46–48]. These topics underscore work being done to study how tumors can alter their immunogenicity and immune evasion tactics, potentially informing CAR T-cell therapies or selection of specific antibodies which can be applied in a personalized manner [49–51]. Revealing additional heterogeneity may refine the selection of adjuvant therapy choices outside of existing prognostic measures (e.g., pTNM staging) depending on the kind of immune reactivity.

### Limitations and Future Directions

Our study, while promising, has several limitations that offer avenues for future research. First, the generalizability of our detection neural network models may be constrained due to the limited sample size of the Lizard dataset used to train the cell detection models, which may not be representative of the broader colorectal tumor population. Inaccurate cell detection may have hampered the predictive performance of the cell-graph neural networks. Additionally, while our cohort of nine samples is large for a Visium study, given its cost, plans are underway to amass a larger, more diverse cohort to bolster the robustness of our findings by accounting for further tumor heterogeneity. As our cohort was restricted to pT3 patients, future work will examine the predictiveness of these algorithms at additional tumor sites and levels of invasiveness, particularly in relation to the tumor-immune microenvironment. Another limitation is that our cell graphs solely relied on local connectivity for information flow, lacking positional embeddings or integration with patch-level information. The slight performance advantage of convolutional neural networks (CNNs) over graph neural networks (GNNs) could be attributed to CNNs’ inherent structural benefits. Future work could explore the benefits of deeper or more extensive cell graphs and the incorporation of single-cell RNA priors for more accurate predictions. While cell regularization and similar strategies might not enhance the ultimate predictive capability for pooled Visium expression, they do bolster performance concerning optimal transport between single-cell profiles assigned to Visium spots using deconvolution methods (like Tangram). Direct model predictions can display notable correlations in gene expression levels across the cell distribution within each spot, even without an input single-cell profile. In the future, these methods could serve as adjustable enhancements or alternatives to Tangram. Inaccurate mapping of single-cell profiles to Visium spots may have also impacted the validity of the associations between single-cell inferred expression and could improve with the adoption of other single-cell spatial mapping methods. Potentially this approach could be used to map single cell profiles, where scRNASeq information has been collected, to their precise locations within slides without the use of Visium as an intermediary. Our approach could also extend to identify other spatially-resolved molecular features, such as protein interactions [52,53]. Validation of our findings using pathological examination of underlying cellular phenomena, through immunohistochemistry and other single-cell level spatial analysis platforms like Xenium/merScope is essential [54,55], as this study serves as a proof of concept. We anticipate applying our models to a larger cohort to identify metastasis and recurrence predictors at a population scale. Furthermore, the trade-off in model accuracy is mitigated by the potential for scalability, allowing for the assessment of a larger number of genes with reasonable predictive performance and statistical power. Overall, our study signifies a crucial step towards improving cancer diagnostics and prognosis by incorporating spatial transcriptomics into histological images, and future efforts will focus on refining these techniques.

Ultimately our findings could shed light on the molecular alterations occurring in these immune cell subsets, which may help identify potential immune evasion mechanisms employed by tumor cells, such as the upregulation of immune checkpoint molecules or the recruitment of immunosuppressive cell populations. By integrating this spatial and molecular information, we can potentially uncover the functional “hotspots” within the tumor microenvironment, where effective immune responses are either promoted or hindered. These functional “hotspots” could ultimately aid in designing immunotherapeutic strategies that aim to reinvigorate the anti-tumor immune response, either by modulating the activity of immune cells in these hotspots or by altering the spatial organization of immune cells within the tumor microenvironment.

## Conclusion

Our investigation into the spatial patterns of T cells, NK cells, and B cells near the primary tumor site and their molecular alterations has potential implications for our understanding of tumor microenvironment dynamics and the broader clinical and immunological context. These patterns can offer valuable insights into the potential for concurrent nodal and/or distant metastasis and serve as indicators of the risk of recurrence and mortality. They provide critical complementary information to traditional lymph node assessment screening programs. Understanding these patterns can significantly influence disease management strategies for patients with colorectal cancer, as the immune response is intrinsically tied to the disease’s pathogenesis and progression. The promise of spatially inferring gene expression patterns from routine histological staining within larger cohorts could vastly enhance our understanding of specific transcriptomic changes occurring within distinct spatial architectures associated with metastasis. Traditionally, conducting highly multiplexed spatial profiling in such cohorts has been cost-prohibitive. However, with the advent of new methods, it is now feasible to explore and analyze spatial transcriptomic data at a previously challenging scale. This advancement opens up unprecedented possibilities for more cost-effectively investigating spatial cell-type specific changes associated with metastasis.

Our study revealed that by considering cells’ histomorphology and spatial relationships, we could effectively predict gene expression patterns across whole slide images and recover local patterns of cellular heterogeneity. Identifying structural cell organizations that exhibited the strongest correlation with the expression of specific genes has the potential to drastically improve our understanding of the tumor-immune microenvironment dynamics and potentially guide personalized treatment plans. Future applications of this method could include predicting response to immunotherapy based on the spatial distribution and expression patterns of immune cells in the tumor microenvironment. In conclusion, our study underscores the potential of integrating spatial transcriptomic information into histological images using feature engineering approaches. It offers a promising direction for enhancing not only the diagnosis and prognosis of cancer but also our broader understanding of the clinical and immunological intricacies of the tumor microenvironments. As the field of spatial transcriptomics advances, we anticipate that this methodology will play an increasingly pivotal role in shaping personalized and precision medicine strategies for cancer treatment.

## Methods

### Data Collection and Preprocessing

The dataset used in this study comprised nine patients with pathologic T Stage-III (pT3) colorectal cancer. Following IRB approval, these patients were selected through a retrospective review of pathology reports from 2016 to 2019. Notably, this cohort is distinct from our initial cohort of four slides, profiled using the original Visium assay without using CytAssist.

Patients were matched based on various criteria such as age, sex, tumor grade, tissue size, mismatch repair/microsatellite instability (MMR/MSI/MSS) status, and tumor site. MSI status was determined through the loss of expression of MLH1 and PMS2 as assessed through immunohistochemistry (there was no loss of expression for MSH2 and MSH6).

The dataset included both patients with concurrent tumor metastasis and patients without metastasis, with equal representation from each group. Patients were carefully selected to ensure a balanced representation across various factors, including tumor site, grade, node status, MSI status, and sex. Tissue blocks were sectioned into 5-micron thick layers. Specific regions of interest within these sections, including epithelium, tumor-invasive front, intratumoral areas, and lymphatics, were annotated by a board-certified GI pathologist. Following annotation, these regions were dissected from the tissue, and subjected to H&E staining, imaging, and Visium profiling at the Pathology Shared Resource at Dartmouth Cancer Center and Single Cell Genomics Core in the Center for Quantitative Biology.

#### Enhanced Pathology Workflow for Visium Profiling

In this study, we utilized the enhanced workflow described in(10)., to profile specimens utilizing the Visium assay. Forv four slides, which corresponded to eight patients, we macrodissected tissue sections from Formalin-Fixed Paraffin-Embedded (FFPE) blocks into 5.5mm by 11mm rectangular segments—precisely delineated by the pathologist in serial Whole Slide Images (WSI)—to isolate specific tissue architectures. To optimize study expenses, rectangular tissue segments from two different patients were juxtaposed at the center of a standard histology slide to form 11mm^2^ capture areas (each corresponding to approximately 14,300, 50-micron Visium spots) and then secured with a coverslip. Within each of these dual-patient capture areas, we maintained an equal representation of metastasis and MSI status and ensured each capture area comprised tissue from similar anatomic sites. For one of these slides, we profiled approximately 5,000 50-micron Visium spots within a 6.5mm by 6.5mm capture area. Our analysis involved a total of five tissue capture areas, representing nine distinct patients. The characteristics of the patients, such as age, sex, and tumor site for these batches, are provided in **Table 2**.

**Table 2:** Patient characteristics. This table details nine unique patients’ profiles across five capture areas. Four of these areas are divided equally between two patients, while one is dedicated to a single patient. Critical patient data, including microsatellite stability status, metastasis occurrence, and tumor site, are provided for each individual.

| Capture Area | Dimensions (mm) | Section Placed on Left Side / Center of Capture Area |  |  |  |  | Section Placed on Right Side of Capture Area |  |  |  |  |
| --- | --- | --- | --- | --- | --- | --- | --- | --- | --- | --- | --- |
|  |  | Age | Sex | Tumor Site | MSI Status | Metastasis | Age | Sex | Tumor Site | MSI Status | Metastasis |
| 1 | 6.5 x 6.5 | 80-85 | M | Left Colon | MSI | yes | — | — | — | — | — |
| 2 | 11 x 11 | 90-95 | F | Hepatic Flexure | MSI | yes | 80-85 | M | Left Colon | MSI | no |
| 3 | 11 x 11 | 85-90 | F | Splenic Flexure | MSS | yes | 75-80 | F | Hepatic Flexure | MSS | no |
| 4 | 11 x 11 | 80-85 | F | Cecum | MSI | yes | 70-75 | M | Cecum | MSI | no |
| 5 | 11 x 11 | 55-60 | F | Left Colon | MSS | yes | 65-70 | M | Sigmoid | MSS | no |

To achieve uniform staining and enhance image quality, we incorporated the CytAssist workflow, which allows Visum profiling of tissues on standard histology slides, enabling the use of pathology department automated staining (Sakura Tissue-Tek Prisma Stainer– Sakura Finetek USA, Inc. 1750 West 214th Street, Torrance, CA 90501) and advanced WSI at 40x resolution (0.25 micron per pixel) via Aperio GT450s to obtain reproducible, high-quality images. Following the preparation of the tissue slides, we employed the Visium assay using the CytAssist technology acording to the manufacturer’s protocol (CG000495) [56,57]. Here, transcriptomic probes are hybridized to the tissue section and ligated to form an amplifiable product. CytAssist facilitates the precise transfer of the ligated probes from the tissue slide to the Visium CytAssist Spatial Gene Expression slide (GEX), which harbors spatially-barcoded capture oligonucleotides, bearing a poly-A sequence complementary to the probes. During transfer, CytAssist images the tissue within the fiducial frame of the GEX slide, allowing for the co-registration of 40x H&E tissue images with Visium ST spots. transferred probes are then extended and amplified prior to sequencing on an Illumina NovaSeq instrument, targeting 50,000 reads/spot. This comprehensive procedure allows for unbiased and gridded profiling of up to 5,000 spots, each containing 1-10 cells, within a 6.5 mm^2^ capture area, and up to 14,300 spots within an 11 mm^2^ capture area. For data processing, we utilized Spaceranger V to align the CytAssist images with the corresponding 40X H&E stains, conduct quality control, and convert the Visium Spatial Transcriptomics (ST) data into a spots x genes expression matrices used for downstream analysis [58].

#### Single Cell Profiling

We utilized the Chromium Flex assay to acquire single-cell RNA-Seq data, specifically from serial sections of patients identified in Capture Areas 2 (left section) and 5 (right section), as detailed in **Table 2**. This method allows for single cell profiling of disaggregated FFPE tissue sections using the same transcriptomic probe set as the Visium assay, thereby revealing the diverse cell types within the tissue. This method was performed according to the manufacturer’s Demonstrated Protocol (CG000606). Data were processed using CellRanger v7.1.0 to generate quality control metrics and a cells by genes expression matrices for downstream processing.

### Preprocessing and Augmentation

#### Selection of Gene Prediction Targets

We curated a list of 1,000 target genes by initially filtering out those not appearing in at least 100 spots per patient. These genes were subsequently ranked based on the fraction of their spatial variance, as determined through SpatialDE analysis. To rectify aberrant gene expression levels, we applied a transformation to both prediction and target gene counts using the expression log(1 + counts).

#### Extraction of Single Cell Imaging Information

Cell detection was performed using the Mask-RCNN framework, which was trained on both the Lizard dataset and our internal dataset [59–61]. The nuclei detection model, available through the public Detectron2 Model Zoo, served as our pre-trained base. This model was further trained on our dataset for up to 5,000 epochs, with training halted upon the observation of overfitting (indicated by a peak in mAP on the validation set). The architecture employed was a Mask RCNN with a Residual Network + Feature Pyramid Network (ResNet+FPN) backbone based on the ResNet-101 model. After training, this cell detection model was systematically applied across each Whole Slide Image (WSI).

#### Image Normalization and Augmentation

The associated image was normalized for each detected cell through standard scaling applied over the image channels. We implemented data augmentation techniques to enhance our dataset, including random rotations (up to 90°) and color jitter adjustments. These augmentations were specifically applied to the images and cell detections cropped around the Visium spots during the training phase.

### Deep Learning to Integrate Information from Localized Cells to Predict Spatial Gene Expression

Cell graph neural networks (CGNN) facilitate the exchange of messages between adjacent cells, enabling the exchange/incorporation of contextual information [62–67]. This approach effectively captures the relationships between different cell populations within the tissue, including tumor cells and surrounding immune and other cell subpopulations. The important difference between this approach and a more general convolutional neural network model is that cells and the relationships between them are modeled explicitly. Leveraging these relationships can enhance the predictive performance of our spatial RNA inference algorithms while providing additional information as to relevant cells for these predictions.

We implemented an end-to-end training strategy that integrates the simultaneous training of a Convolutional Neural Network (CNN) and a Graph Neural Network (GNN). The CNN is designed to extract cell-level features from histological images, while the GNN contextualizes these features by incorporating information from neighboring cells during the model-fitting process. This stands in contrast to previous cell-level neural network models, which typically employ a two-stage approach. In these two-stage models, features are initially extracted using a CNN, and these extracted features serve as input to train the GNN in a separate, subsequent step. Such a two-stage paradigm can potentially lead to suboptimal results, as the CNN’s parameters are fixed before the GNN training begins, thereby limiting the CNN’s ability to adapt based on the contextual information that the GNN could provide. Our end-to-end approach aims to harmonize the feature extraction and contextualization processes, enabling the CNN to learn cell-level features that are more effectively contextualized through iterative, integrated training with the GNN (**Figure 1,2**).

The backbone of the model is a four-layer graph attention network (GAT) [68,69], which uses self-attention mechanisms to update the representation of each cell with the information of its neighbors. We extract nodal attributes from detected cells using a ResNet-50 model, which is trained jointly with the graph attention layers. The Euclidean distances between the spatial locations of detected cells are used to form k-nearest-neighbor cell graphs (k=6, determined through a sensitivity analysis), connecting cells based on their spatial proximity. Patients within the same Visium capture area were divided according to pathologist-annotated segmentation masks– we ensured that cell graphs did not overlap across patients. The model updates cell-level embeddings through information sharing up to four neighbors away, facilitated by message passing between nodes across four graph attention layers, each mapping cells to 512-dimensional numerical vectors. Final node embeddings pass through a linear layer, producing a vector representing each gene’s relative pseudocount-transformed expression for each cell. Cells corresponding to the same Visium spot are then aggregated through global sum pooling to predict expression at the Visium spot. This is then compared to the pseudocount-transformed ground-truth Visium data with mean squared error.

### Comparison of Cell-Graph Neural Network Regularization Strategies

In addition to evaluating the congruence between ground truth and predicted expression at the spot level, we explored the following methodological variations:

1. **Vanilla Supervised Learning Objective:** This baseline approach focuses solely on the supervised learning objective, serving as a reference for evaluating the potential gains from additional regularization strategies.
2. **Incorporating Graph Contrastive Learning:** This approach introduces a self-supervised regularization term that encourages the model to learn embeddings through the comparison of augmented viewpoints of the same cell graph / Visium spot to different cell-graphs / Visium spot. This can enhance the model’s sensitivity to spatial patterns in the data, potentially improving its predictive accuracy for spatial transcriptomics patterns.
3. **Incorporating Single-Cell RNA-Seq Penalization through Optimal Transport:** This strategy introduces a penalty term that encourages the model to align cell-level histological features more closely with corresponding single-cell RNA-Seq data. By leveraging optimal transport theory, this term effectively “guides” the model towards a solution where the spatial patterns inferred from histology are maximally consistent with independent single-cell RNA-Seq measurements, thereby enhancing the biological validity of the model’s predictions.
4. **Combining Graph Contrastive Learning and Single-Cell Penalization:** This approach synergistically combines both the graph contrastive learning and the single-cell RNA-Seq penalization strategies, aiming to leverage the benefits of both spatial context awareness and alignment with single-cell RNA-Seq data. This dual-regularization strategy is designed to promote a model that is both sensitive to spatial patterns and tightly aligned with independent molecular measurements, potentially offering a balance between spatial sensitivity and biological validity.

### Graph Contrastive Learning

Using the PyGCL package, graph contrastive learning was implemented through augmentations to random cell positions in the nearest neighbor graph construction, dropping edges with a probability of 0.1, and masking out features with a probability of 0.3. Graph contrastive learning is a form of self-supervised learning that can improve the generalizability and robustness of graphs [70–72]. By intentionally adding noise to the training cell graphs and comparing these representations at different Visium spots, we aimed to improve the model’s generalizability when tested on held-out data by forcing it to learn a stable, robust, foundational representation.

### Incorporating Single-Cell Expression

For two of our patients in the study, we obtained corresponding single-cell RNA-Seq data, which were selected to be representative across sex, microsatellite instability, and metastasis status. Due to budget constraints, we were unable to acquire matching single-cell data for all the slides, a limitation we aim to address in future work. Our approach involves the integration of single-cell RNA-Seq data as a form of model regularization. By encouraging the predictions derived from histological images of individual cells to align closely with the corresponding true single-cell expression profiles, we aim to enhance the interpretability of our models through more consistent and biologically meaningful cellular information. This, in turn, is expected to improve the model’s generalizability. The objective is to develop a nuanced understanding of cell type populations at specific spatial locations while maintaining fidelity to the ground-truth expression data sampled through the Visium platform. By fine-tuning the alignment between these profiles, we strive to increase the likelihood that our predictions accurately reflect the true cellular composition at each spatial location. In contrast to previous attempts to integrate single-cell information with histology, where spatial transcriptomics data were not employed to guide the spatial mapping of single-cell information, our approach uniquely capitalizes on the spatial context offered by the spatial transcriptomics data. Specifically, we align single-cell profiles with individual Visium spots, thereby avoiding spurious cell assignments that can arise when attempting to map single-cell information across entire whole-slide images without the benefit of such spatial guidance.

We initiated our analysis by mapping scRNA profiles to Visium spots using the Tangram tool [73], and we selected the top *k* most likely cells to be assigned to each spot, where *k* represents the number of detected cells in that spot. In the context of our CGNN model, which aims to predict log-expression levels for individual cells, it was critical to assess the alignment between these predictions and the assigned single-cell profiles. Originally, we employed a dynamic matching approach wherein the pool of cells assigned to each spot was paired with our detected cells based on the Euclidean distance between their predicted and observed expression profiles. This matching was formulated as a variant of the linear sum assignment problem, solved using the Hungarian algorithm. This algorithm established the optimal one-to-one correspondence between detected cells and their closest-matching assigned scRNA cells, thereby minimizing the Euclidean distance between them. We applied a mean squared error (MSE) loss across the log-transformed expression values to penalize discrepancies between these matched profiles. However, following internal comparisons, we shifted our approach to leverage the Wasserstein loss as a more effective metric for aligning our predictive single-cell expression profiles with the true expression profiles derived from scRNA data. The Wasserstein distance, a loss metric formulated on principles from the theory of optimal transport [74–77], quantifies the minimum cost required to transform one distribution into another, effectively capturing the overall distributional differences between predicted and ground truth single-cell expression profiles. This approach offers more flexible comparisons, as it can accommodate shifts in the distribution and discrepancies in the shape of the distributions without necessitating a direct one-to-one matching between predicted and observed cells.

Our internal comparisons revealed that the Wasserstein loss provided more precise and interpretable mappings between detected and observed cellular profiles, solidifying it as our chosen approach. The Wasserstein loss was computed for each Visium spot and used to compare the effectiveness of different regularization methods, measuring alignment between predicted and ground truth single-cell expression.

### Comparison to Convolutional Neural Network Approaches

The CGNN approaches were compared to patch-based convolutional neural network methodologies deemed highly predictive from previous works– namely the InceptionV3 neural network. The Inceptionv3 network can capture features at various receptive fields. This model was trained on images of tissue patches encompassing multiple cells inclusive of surrounding matrixed tissue architecture. We initialize the model with ImageNet weights (with the final layer truncated) on the same dataset. We apply the same visual transformations as for the cell embeddings, (standard scaling, color jitter, random 90° rotations).

### Training and Validation

CGNN models were implemented with the torch-geometric Python package [78], which extends the PyTorch machine-learning framework. We use PyGCL [72] to apply graph augmentations. CGNN were trained using the Adam optimizer [79] with a learning rate of 0.0001 on one Nvidia V100 GPU with Dartmouth Research Computing, quickly converging after two epochs. Similarly, the CNN model was trained for around 100000 iterations on a Nvidia V100 GPU.

The final performances of these models were compared using leave-one-patient-out cross-validation. Statistics are reported with the Spearman correlation coefficients. We also sought to assess the performance of predicting dichotomized gene expression (low/high), a binary classification problem– this was accomplished by dichotomizing expression according to [30], used to calculate the area under the receiver operating characteristic curve (AUROC) as another performance measure. Models were trained and then cross-validated using all capture areas except one, reserved for testing, and repeated for all cross-validation folds. Performance statistics were generated for each cross-validation fold, including Spearman’s correlation coefficients and area under the receiver operating characteristic curves (AUROCs) by gene. The results were then averaged across all folds to assess the best-performing model on a gene-specific basis. We calculated 95% confidence intervals for all performance statistics, reported using 1000 sample non-parametric bootstrapping.

### Model Interpretation through Single Cell Attributions, Gene Embeddings and Pathway Analysis

#### Single-cell attributions

We visually inspected the CGNN’s ability to reconstruct single-cell gene expression based on the aggregate neighborhood’s gene expression. Since the CGNN calculates the overall expression of a Visium spot by summing predictions for individual cells, we hypothesized that the predictions for each cell would reflect their relative contribution to the spot’s expression. We believed that the model would learn consistent and meaningful gene expression counts due to the large amount of data and our aggregation method.

Operating under this assumption, we generated *attribution maps* representing each cell’s expression for the 1000 genes across the slide. These multidimensional maps provided insights into the expression patterns of individual cells, even though they were not explicitly labeled for that purpose. To assess the validity of these attribution maps, we collaborated with a pathologist for their expert evaluation. However, we acknowledge that further detailed analysis is needed in future work to delve deeper into their significance and accuracy. To complement these attribution maps, we also generated analogous *explanation maps*, which utilized the GNNExplainer algorithm to assign importance scores to specific cells based on their relevance for predictions of specific genes [80].

#### Gene embeddings

Similar to our previous work, we sought to intuitively understand how well each approach could recapitulate the relationships between the Visium spots. This recapitulation was accomplished by applying Uniform Manifold Approximation and Projection (UMAP) to each predicted expression profile [81]. Each method’s predicted and actual gene expressions were aligned and clustered using the AlignedUMAP method. Clusters determined by running HDBSCAN [82] on the ground truth expression data were overlaid on top of the UMAP plots for the other methods. To do this, we created HDBSCAN clusters with a minimum size of 3 across the log-transformed ground truth Visium data. Then, we annotated each of our prediction points with the corresponding HDBSCAN cluster of the ground truth and performed an aligned UMAP, jointly minimizing the distance between similar expressions in the embedding space and between paired ground truth and true locations. In addition, we annotated our histology images with the HBDSCAN clusters to interpret the tissue type of origin for each point. The visual similarity of clustering patterns would indicate an overall similarity of the predicted and ground truth expression.

#### Pathway analysis

Pathway analyses were performed to assess the ability of the methods to capture broader biological phenomena. Pathway analyses were accomplished using two separate methods: 1) aggregating the Spearman correlation and AUROC statistics across genes associated with pathways identified from the MSigDB Hallmarks gene set, and 2) evaluating the enrichment of the highest genes as ranked using their performance statistics, utilizing enrichR, which employs a modified Fisher’s exact test. By examining the average performance across pathway analysis and overlap tests for the top-performing genes, we can gain insights into which biological phenomena each method effectively represents.

## Declarations

### Ethics approval and consent to participate

Human Research Protection Program IRB of Dartmouth Health gave ethical approval for this work.

### Availability of data and materials

Access to manuscript data is limited due to patient privacy concerns. All data produced in the present study are available upon reasonable request. Requests should be directed to senior author Dr. Joshua Levy.

## Acknowledgements

This study was carried out in the Genomics and Molecular Biology Shared Resource (GMBSR) at Dartmouth which is supported by NCI Cancer Center Support Grant 5P30CA023108 and NIH S10 (1S10OD030242) awards. Additionally, single cell genomics projects should include the following text (or similar): “Single cell studies were conducted through the Dartmouth Center for Quantitative Biology in collaboration with the GMBSR with support from NIGMS (P20GM130454) and NIH S10 (S10OD025235) awards.

## Supplementary Material

**Supplementary Table 1:** Spearman correlation scores for top performing genes from each CGNN method.

| Vanilla |  |  | Graph Contrastive Pretraining |  | Single-Cell Regularization |  | Both |  |
| --- | --- | --- | --- | --- | --- | --- | --- | --- |
|  | Gene | R | Gene | R | Gene | R | Gene | R |
| 1 | LGALS3BP | 0.484 | S100A6 | 0.575 | FXYD3 | 0.624 | TFF3 | 0.643 |
| 2 | KRT8 | 0.443 | FXYD3 | 0.568 | LGALS4 | 0.590 | KRT8 | 0.638 |
| 3 | SPINT2 | 0.425 | TMSB10 | 0.566 | KRT8 | 0.590 | S100A6 | 0.634 |
| 4 | MYL6 | 0.412 | TMSB4X | 0.561 | LGALS3 | 0.574 | FXYD3 | 0.625 |
| 5 | KLF5 | 0.404 | TFF3 | 0.551 | MYH14 | 0.567 | PHGR1 | 0.602 |
| 6 | S100A6 | 0.403 | MT-ND4L | 0.550 | CLDN4 | 0.560 | PIGR | 0.595 |
| 7 | JUN | 0.392 | PIGR | 0.548 | MISP | 0.556 | LGALS3 | 0.592 |
| 8 | ETS2 | 0.390 | KRT8 | 0.546 | S100A6 | 0.541 | ELF3 | 0.592 |
| 9 | CSRP1 | 0.390 | LGALS3 | 0.543 | PHGR1 | 0.540 | CLDN7 | 0.590 |
| 10 | ACTB | 0.389 | MT-CO3 | 0.537 | TFF3 | 0.535 | KLF5 | 0.589 |
| 11 | PHGR1 | 0.385 | LGALS4 | 0.535 | CLDN7 | 0.534 | CD24 | 0.586 |
| 12 | S100A10 | 0.384 | ITM2C | 0.534 | CD24 | 0.530 | EPCAM | 0.585 |
| 13 | ZFP36 | 0.377 | MT-CO2 | 0.530 | CD9 | 0.521 | LGALS4 | 0.583 |
| 14 | PERP | 0.377 | PHGR1 | 0.526 | EPCAM | 0.519 | CLDN4 | 0.583 |
| 15 | IGFBP4 | 0.373 | MT-ND4 | 0.510 | TMSB10 | 0.505 | FCGBP | 0.580 |
| 16 | COL1A1 | 0.370 | EPCAM | 0.501 | PIGR | 0.502 | MYH14 | 0.580 |
| 17 | FLNB | 0.367 | CLDN4 | 0.501 | SPINT2 | 0.501 | TMSB10 | 0.579 |
| 18 | SDC4 | 0.363 | MT-ND6 | 0.494 | REG4 | 0.499 | TMSB4X | 0.567 |
| 19 | CLDN7 | 0.354 | STARD10 | 0.490 | KLF5 | 0.496 | MISP | 0.565 |
| 20 | LMNA | 0.348 | LGALS3BP | 0.490 | ANXA2 | 0.491 | STARD10 | 0.564 |
| 21 | CDC42EP5 | 0.346 | CLDN7 | 0.489 | PPP1R1B | 0.481 | CD9 | 0.559 |
| 22 | CANT1 | 0.344 | MISP | 0.479 | IFI27 | 0.479 | LGALS3BP | 0.554 |
| 23 | PYGB | 0.337 | CD24 | 0.477 | STARD10 | 0.479 | SPINT2 | 0.551 |
| 24 | MYC | 0.334 | MT-CYB | 0.467 | MLEC | 0.479 | MUC13 | 0.549 |
| 25 | GMDS | 0.327 | ACTB | 0.457 | ELF3 | 0.475 | SDC4 | 0.542 |

**Supplementary Figure 1:**
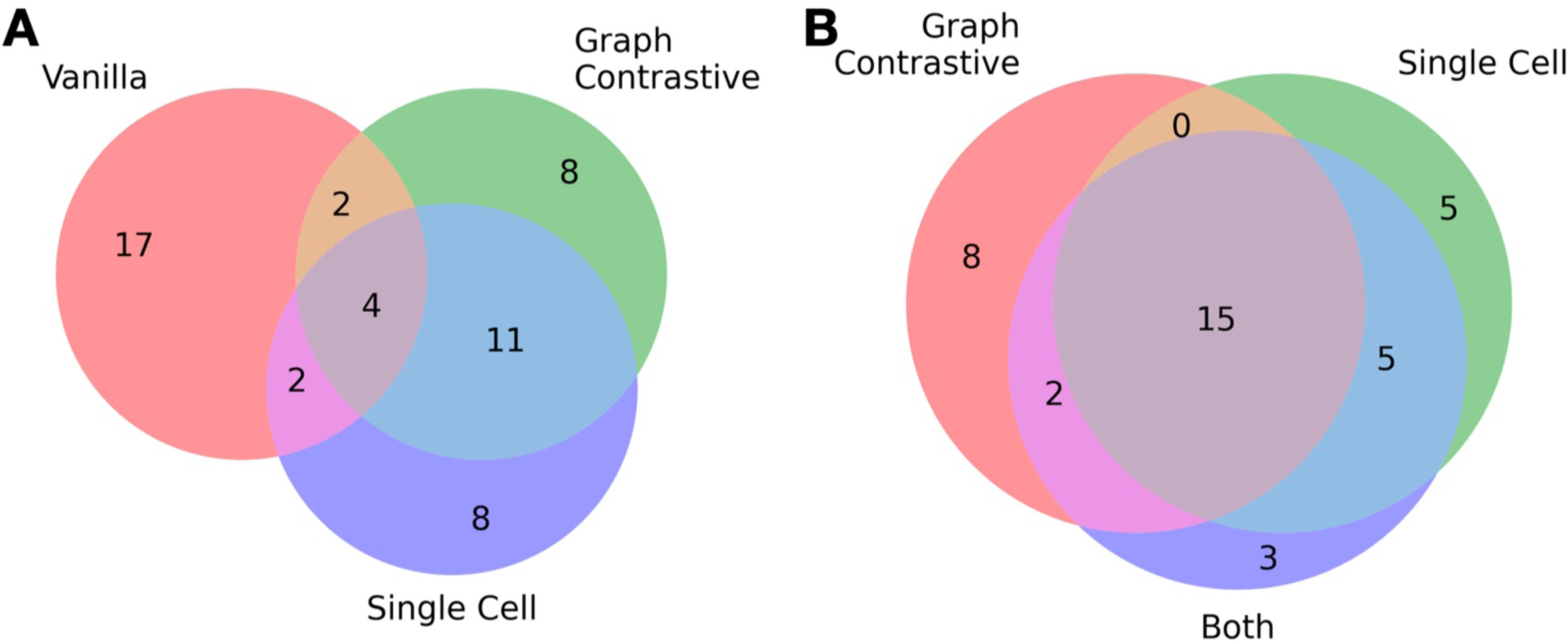
Comparative analysis of top performing genes used in different penalization and pretraining methods for cell graph neural networks predicting spot-level gene expression. **A)** Left Venn Diagram: Shows the overlaps between genes selected using the “Vanilla”, “Graph Contrastive”, and “Single Cell” penalization methods. Each circle represents a distinct set of genes, with overlapping regions indicating shared genes between methods. **B)** Right Venn Diagram: Represents the overlaps between genes selected using the “Graph Contrastive”, “Single Cell”, and “Both” (a combination of methods) penalization/pretraining approaches. Overlaps indicate genes that are common across multiple methods, highlighting the consensus and divergence in gene selection across these strategies. Numbers within each segment of the diagrams indicate the count of genes unique to or shared by the corresponding methods.

**Supplementary Figure 2:**
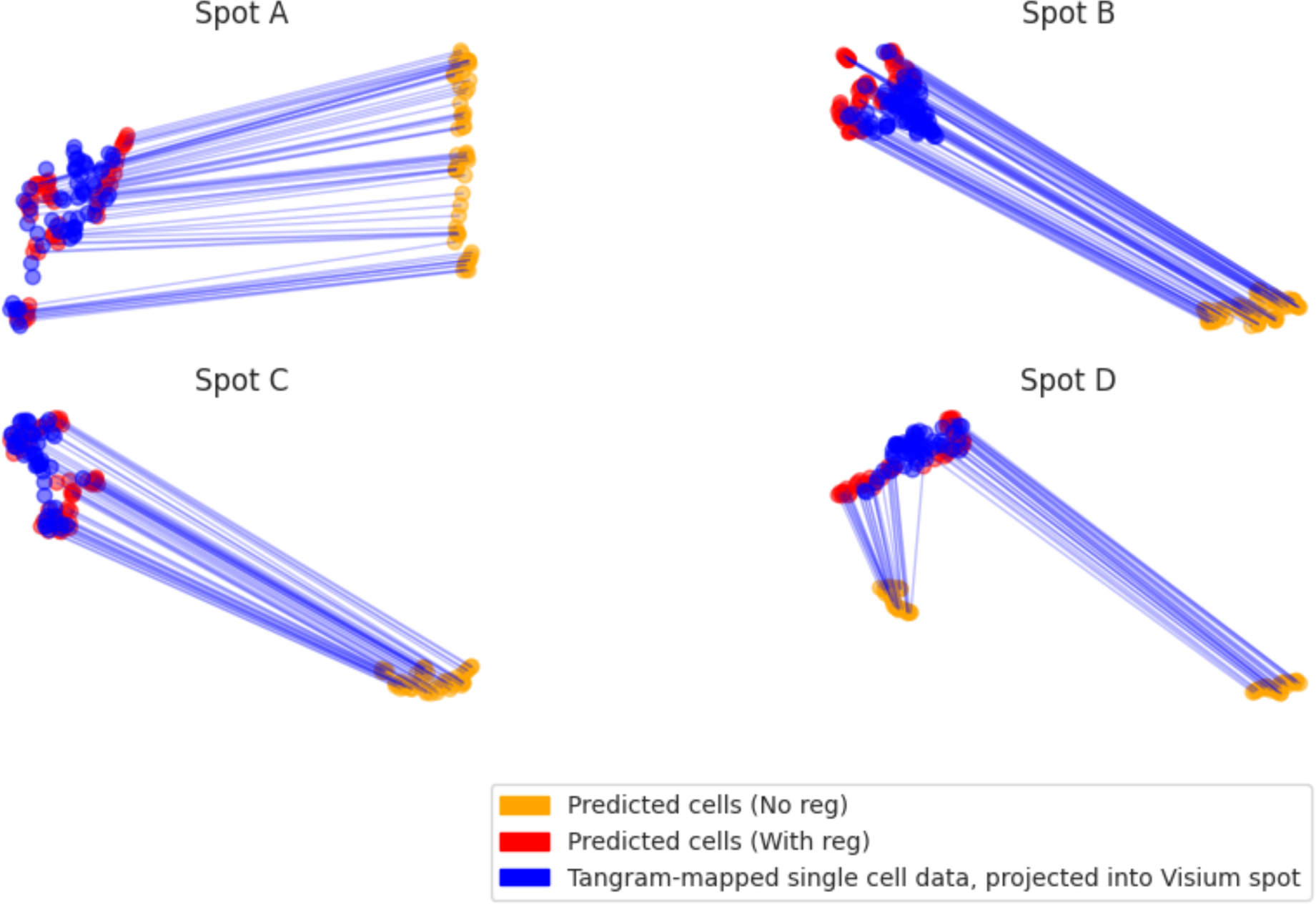
UMAP Embedding of Predicted vs. Assigned Cell Expressions. This figure showcases UMAP embeddings contrasting predicted cell expressions against expressions of cells designated to the Visium spot via the Tangram optimal matching algorithm. The red embeddings showcase model outputs with regularization, blue embeddings correspond to the original ground truth from single cell RNA-Seq, and yellow embeddings represent predicted single cell outputs without regularization for the same spot. Evidently, cell regularization enhances the overlap of predicted and assigned gene expressions. Performance assessment relies on the Earth Mover’s Distance (EMD) between a held-out single-cell profile designated via Tangram and predicted expressions on an isolated slide. Distances between cells employ the Euclidean metric, while optimal matchings leverage the Hungarian algorithm via the “scipy.optimize.linear_sum_assignment” function. Notably, post-regularization, the EMD decreases from 0.2113 ± 0.0018 to 0.1473 ± 0.0018.

**Supplementary Figure 3:**
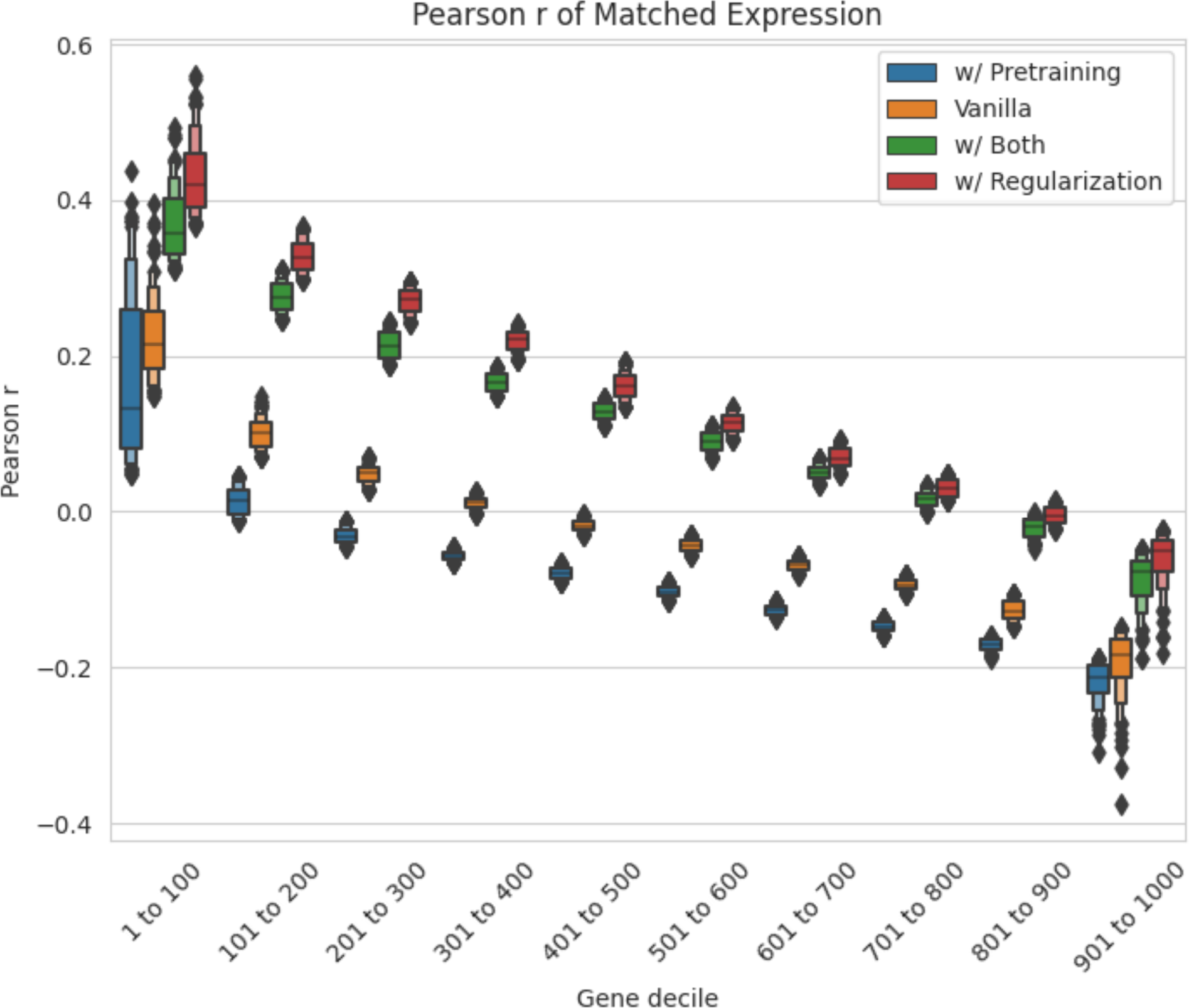
Correlation of predicted versus true single cell log-expression for different deciles of genes, ranked based on performance at the Visium spot level.

**Supplementary Figure 4:**
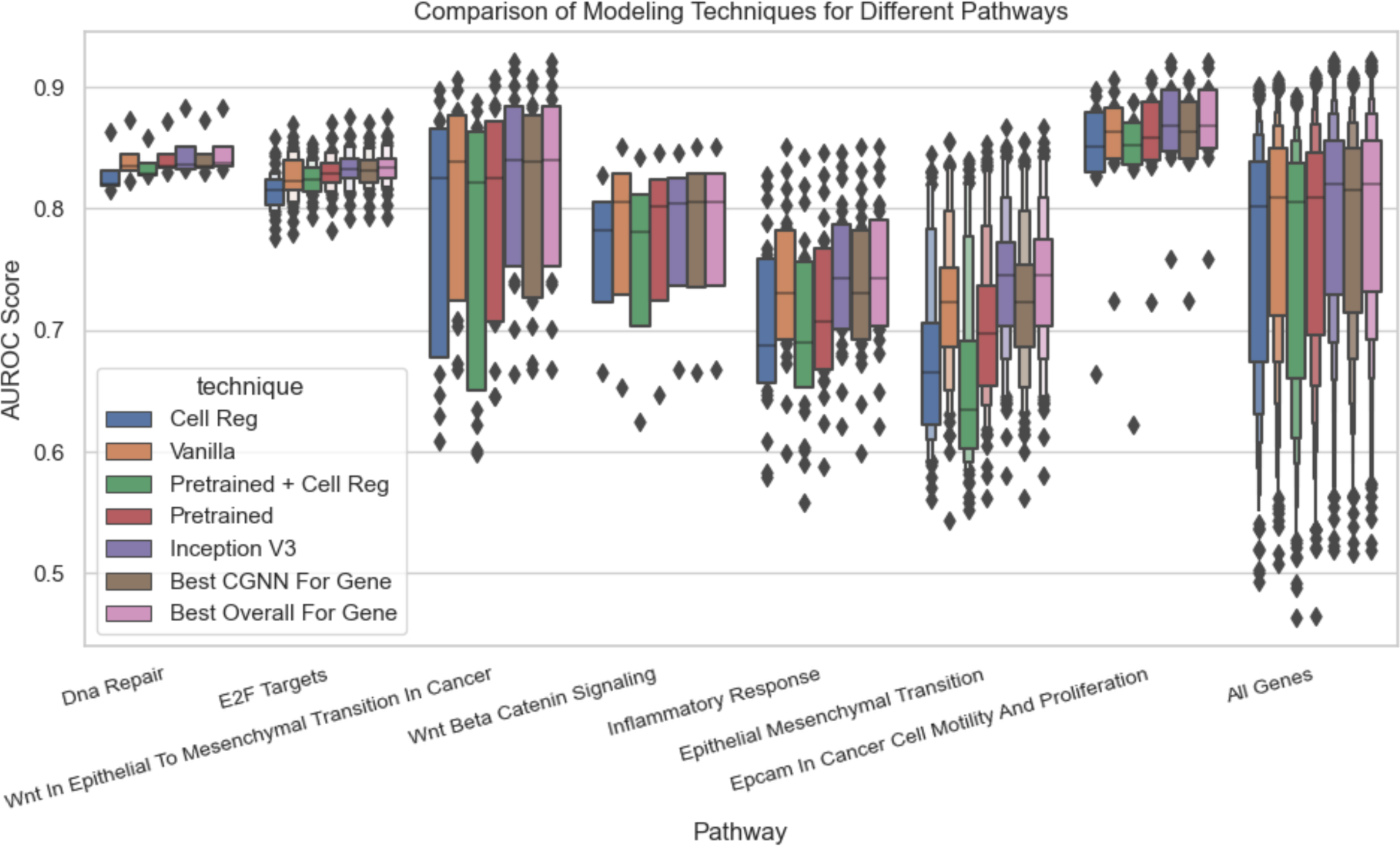
Boxplot of AUROC scores for selected pathways for all modeling approaches.

**Supplementary Table 2:**
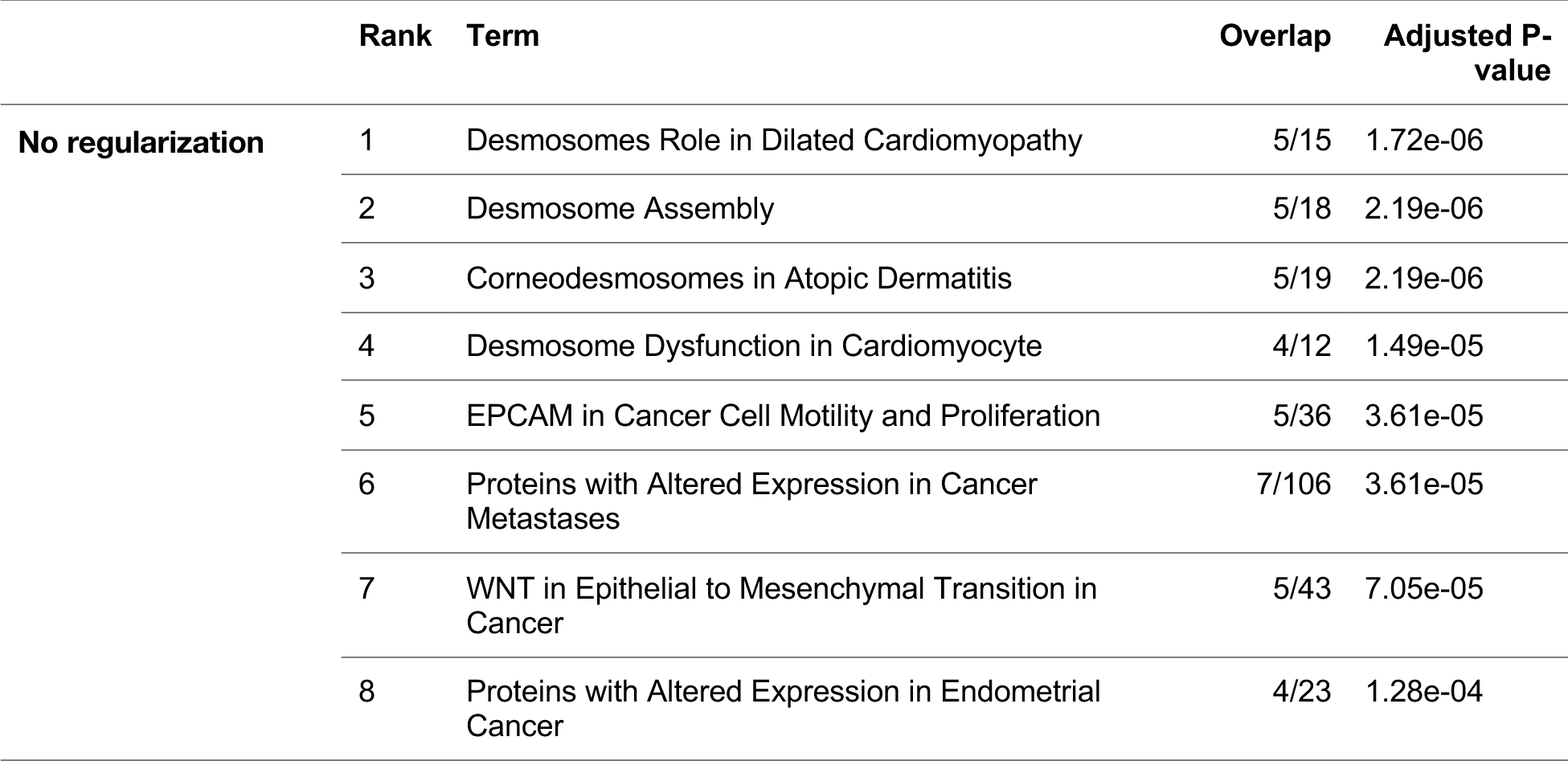
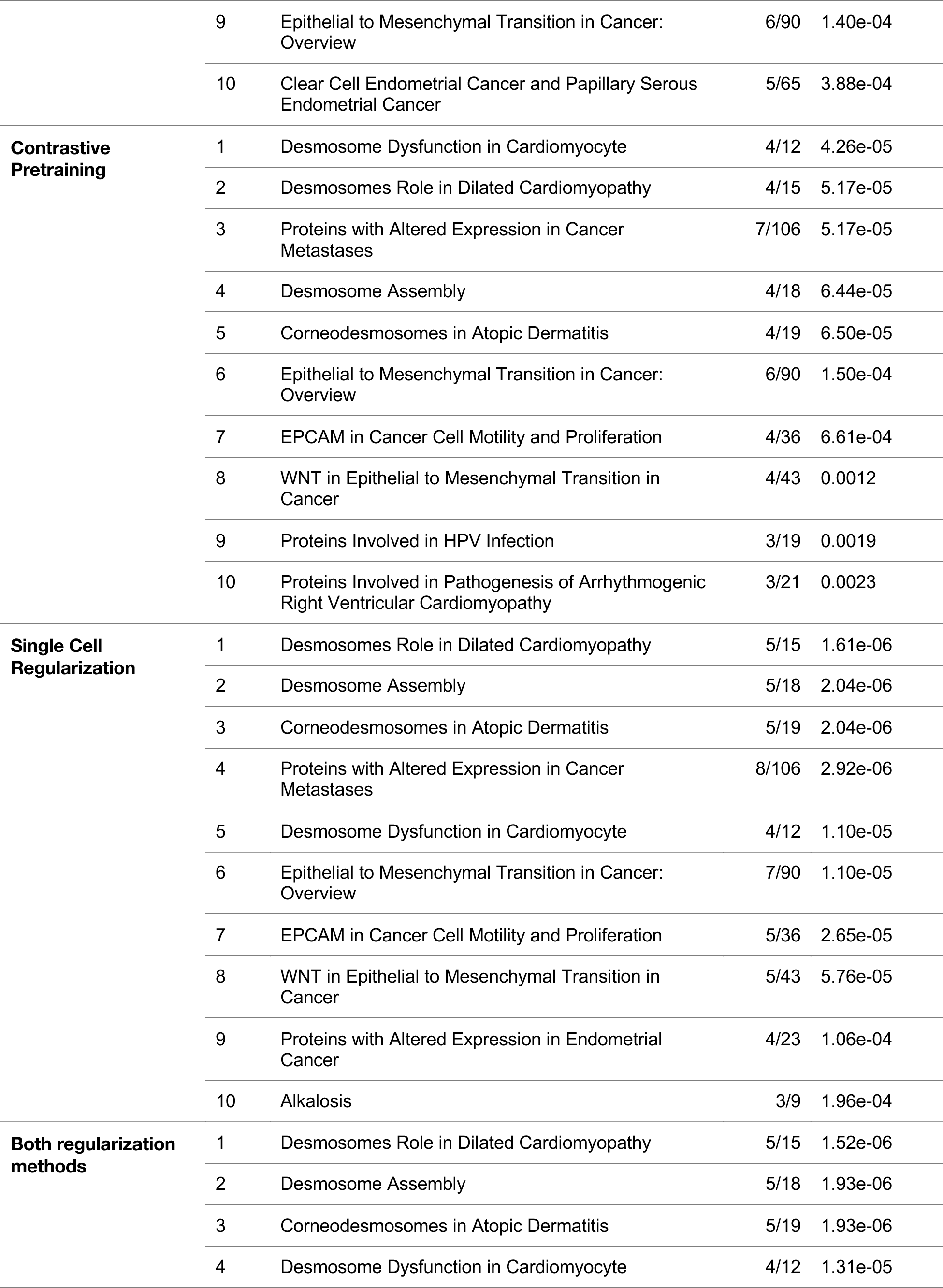
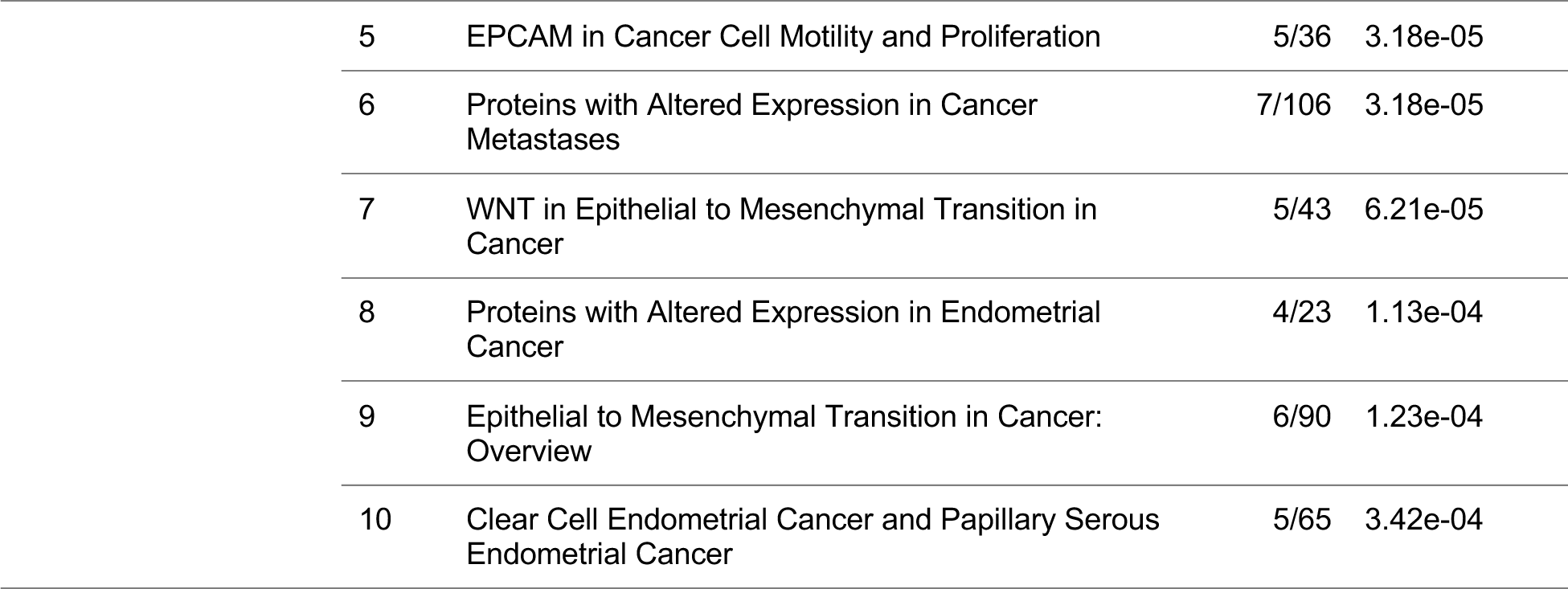
Top 10 pathways ranked by adjusted p-value, sourced from the Elsevier Pathway Collection via Enricher, for each cell-level modeling strategy.

